# Beyond the Diagnostic Label: Data-Driven Characterisation of Cognitive and Mental Health Profiles in a Transdiagnostic Sample of Neurodivergent Adults

**DOI:** 10.1101/2025.09.01.25334849

**Authors:** Karin Madericova, Joel B Talcott

**Author notes:** Corresponding Author Joel B Talcott, Institute of Health & Neurodevelopment College of Health and Life Sciences Aston University, Birmingham, B4 7ET.

## Abstract

Neurodiverse diagnoses (NDs) have been largely conceptualised as distinct categories with clear-cut diagnostic boundaries. Converging research evidence, however, is showing that this conventional approach inadequately captures the individual variation and overlaps that are commonly manifest. This study aimed to build upon emerging quantitative research frameworks for understanding diagnostic complexity, through adopting a transdiagnostic dimensional approach toward characterising the cognitive and mental health dimensions associated with NDs. A cohort of 175 adults, who had been referred for assessment and/or sought help from community support services in the UK for suspected or previously diagnosed neurodiverse conditions, were recruited using convenience sampling. Cognitive domains of executive function, language, attention, processing speed and memory were measured using the National Institute of Health (NIH) Cognition Toolbox*©*. A mental health battery consisting of the Extended Strengths and Weaknesses Assessment of Normal Behaviour (E-SWAN), Strengths and Difficulties Questionnaire (SDQ) and Emotion Regulation Skills Questionnaire (ERSQ) was also administered to provide proxy measures of the mental health. Cluster analyses revealed that the sample of neurodivergent adults was best represented by three data-driven clusters, none of which mapped coherently onto traditional diagnostic labels. These transdiagnostic groups were best characterised by a combination of strengths and weaknesses along two multivariate dimensions: 1) cognitive flexibility and processing speed, and 2) language, memory, emotional wellbeing and behavioural regulation. Overall, this study provides novel, data-driven evidence to support the emerging multivariate, dimensional structure of NDs, and highlights several component processes which contribute to the complexity associated with ND outcomes in adults. Such findings may be used to inform future assessment and/or support strategies for neurodivergent adults beyond those framed by strictly categorical approaches.

## Introduction

### Neurodiverse conditions

Neurodiverse diagnoses (NDs) are behaviourally complex conditions associated with differences in the development of the brain throughout the lifespan. Altered developmental trajectories can lead to individual difficulties in an array of functional domains, including cognitive and affective processing, behaviour and/or communication, with the extent of such difficulties ranging from mild to severe. According to the Diagnostic and Statistical Manual of Mental Disorders (5^th^ ed.; DSM-5; American Psychiatric Association, 2013), NDs encompass conditions such as Autism Spectrum Disorder (ASD), Attention-Deficit/Hyperactivity Disorder (ADHD), or Specific Learning Difficulties (SLD, e.g., dyslexia). The prevalence rates of NDs vary, with a British coalition of charities concerned with NDs (Embracing Complexity) recently suggesting that up to 10% of the UK population may have lived experience of a neurodiverse condition (Alexander et al., 2021).

NDs are among the most frequently diagnosed paediatric conditions, with many of these conditions persisting throughout the lifespan (Cheung et al., 2015; Farah et al., 2021; Orm et al., 2022). Some NDs may not manifest until later in life, while the behavioural manifestations of other NDs are likely to change substantially across development (Caye et al., 2016; Taylor & Seltzer, 2010). Developmental differences may affect a number of functional outcomes, such as social interactions or academic attainment (Duff et al., 2023; McKay et al., 2023). The nature, extent and impact of such outcomes are often dependent on the different ways in which developmental differences manifest across individuals. For example, if an individual with a diagnosis of developmental dyslexia also displays characteristics associated with ADHD (e.g., difficulties with attentional allocation and control), their academic performance is more likely to be affected compared to an individual with dyslexia without co-occurring ADHD, resulting from their more complex and presumed interacting array of cognitive difficulties (Germanò et al., 2010; Willcutt et al., 2001). NDs are highly complex, with characteristic overlaps between diagnoses and high levels of individual variation in their respective aetiologies, characteristics and functional outcomes. Research has shown high co-occurrence rates between many of the most prevalent NDs (e.g., 50-70% of autistic individuals would also receive a formal diagnosis of ADHD; Rong et al., 2021), while weaknesses in specific cognitive domains may also be shared (e.g., processing speed difficulties observed in both dyslexia and ADHD; Willcutt et al., 2010). Furthermore, no two individuals with a given diagnosis will typically present with identical profiles of difficulties, with research providing evidence of substantial within-diagnosis heterogeneity across multiple functional domains, including cognitive and affective processing (Mostert et al., 2018; Yue et al., 2022). Such complexities in the nature, scope and impact of developmental difficulties have significant implications for the way that identification, diagnosis and support of such conditions is framed, both in theory and within application of gold-standard diagnostic structures.

### The Categorical Approach

NDs are currently diagnosed through a variety of educational and/or medical service pathways, each of which typically employs assessment measures derived from traditional diagnostic systems. The DSM-5 has been the most influential framework for diagnosing a range of NDs, embedded in clinical practice, research and therapeutic guidance. For example, the standardisation of diagnoses across different clinicians and treatment providers has enabled many individuals to receive appropriate support. However, there is an emerging debate around the extent to which such diagnostic nosology reflects the complex, multivariate nature of NDs (e.g., Insel et al., 2010; Kotov et al., 2017). For instance, in many cases, the DSM-5 fails to acknowledge the variability within and overlaps between presumed discrete diagnostic categories, as formal diagnoses most often follow strict inclusionary and exclusionary criteria. Although the DSM-5 has attempted to redress this limitation, for example, by reconceptualising autism as a spectrum along which individuals may vary in their manifest traits (APA, 2013), many NDs are still predominantly conceptualised as distinct and invariant categories.

In summary, there are a range of issues with the current diagnostic hegemony. First, the rigidity of current diagnostic criteria inherently excludes many individuals with milder difficulties that do not reach diagnostic thresholds, thus limiting access to support, regardless of the significant impact that such difficulties may have on individual lived experience (Astle et al., 2022). Second, current diagnostic classification systems predominantly focus on the behavioural manifestations of NDs characteristic of childhood, meaning that existing diagnostic criteria may not be well aligned with the variable presentations of NDs observed in adulthood. As a result, it may be more difficult for individuals to receive an adequate diagnosis at later stages in development. This is an emergent issue in the context of the increasing number of adults being referred for assessments and/or seeking help for NDs at present. For instance, the United Kingdom has recently observed a 20-fold increase in adult ADHD diagnoses (McKechnie et al., 2023), emphasising the need for more comprehensive procedures for screening and diagnosing NDs across development.

Further, diagnostic categories have historically shaped research design, with many ‘case-control’ studies recruiting participants based on the presence or absence of a particular diagnosis (Astle et al., 2022). This is problematic as, in reality, only a small proportion of individuals have a ‘pure’ diagnosis of a single ND, with research suggesting that a high percentage of neurodivergent individuals will present with at least one co-occurring condition (e.g., Mohammadi et al., 2019; Kadesjö & Gillberg, 2001; Margari et al., 2013). Consequently, study samples may not accurately represent the range of neurodiversity within the population. Additionally, individuals with subclinical presentations of NDs have the capacity to enrich research with valuable information on the wide range of abilities displayed along the spectrum, yet they are often excluded from research, limiting the advances that can be made in neurodiversity research (Astle & Fletcher-Watson, 2020).

Moreover, many ‘gold standard’ assessment tools employed in the majority of developmental research have been designed to produce dichotomous representations of individual abilities (i.e., ‘typical’ versus ‘atypical’). For instance, the Conners Comprehensive Behaviour Rating Scale (CBRS; Conners, 2008) is widely used by clinical practitioners to assist in the diagnosis of ADHD and for research purposes. The items in this questionnaire, like many other assessment tools, have been directly derived from the DSM-5 diagnostic criteria for ADHD, and although the measure covers various domains of traits associated with the diagnosis, it focuses solely on maladaptive behaviours, without taking into consideration individual strengths. In order to gain a more comprehensive understanding of NDs, it is essential to begin to assess the full range of behaviours displayed by neurodivergent individuals, as opposed to reinforcing the reductionist view that each diagnosis may be traced back to a simple ‘core deficit’ in neurocognitive functioning (Astle & Fletcher-Watson, 2020; Astle et al., 2022).

### The Transdiagnostic Approach

The transdiagnostic dimensional approach promotes a shift away from conventional study designs to more inclusive sampling strategies, novel data collection methods and exploratory data analysis techniques that enable more accurate, data-driven theoretical accounts of NDs. Under this approach, NDs are reconceptualised as a multidimensional space, where neurodevelopment is studied in terms of several dimensions that fall along a continuum, ranging from typical to atypical functioning (Astle et al., 2022). Adopting this approach enables researchers to study developmental difficulties that cut across diagnostic boundaries, as opposed to solely focusing on core characteristics that are presumed to underpin specific diagnostic categories (Dalgleish et al., 2020).

Transdiagnostic recruitment involves the relaxation of strict inclusionary criteria with the aim to capture heterogeneity through sampling a broader population of individuals that vary in the extent and nature of their developmental difficulties (Astle et al., 2022). For instance, participants who do not conform to standard criteria for any specific diagnostic labels, as well as individuals with formal diagnoses, and/or those with more complex profiles of co-occurring conditions may be included in research. Further, traditional tasks are replaced with dimensional measures that enable for the investigation of functions that may not typically be associated with a particular diagnostic label, but may help to better characterise a given difficulty (Astle & Fletcher-Watson, 2020). For example, emotion dysregulation is not included in the diagnostic criteria for any ND, yet it has been established as a transdiagnostic mechanism that has the capacity to predict the severity of co-occurring mental health difficulties in individuals with various NDs, above and beyond specific diagnostic criteria (Cai et al., 2021). Dimensional measures assess relative strengths *and* weaknesses in an area of functioning, thus spanning the whole spectrum of abilities, as opposed to focusing solely on the clinically-relevant trait range – such assessments allow us to reliably and meaningfully distinguish individuals from one another even in subclinical parts of the population, making them well suited for use with transdiagnostic samples (Alexander et al., 2020).

The use of more heterogeneous samples and broader assessment measures naturally produces multivariate data, where data-reduction methods can be used to generate simplified models of such large, varied datasets. For example, cluster analysis may be conducted to identify subgroups of individuals based on their similarities and differences in specific characteristics within a given sample (Rivard et al., 2023; Astle et al., 2022). Importantly, the subgroups (i.e., clusters) identified from dimensional measures are not based on a hypothesis or any predefined grouping criteria (e.g., diagnostic labels), but are fully data-driven, enabling the exploration of symptom profiles that may transcend diagnostic categories (Astle et al., 2022).

Such flexible data-driven accounts of NDs have the capacity to capture shifts in group membership (and thus developmental change) over time, enabling the characterisation, tracking and predicting of individual wellbeing across development (Rivard et al., 2023). Using such techniques, recent transdiagnostic studies have shown that individual profiles of various developmental difficulties may map poorly onto traditional diagnostic labels (e.g., Mareva et al., 2023; 2024), and supporting the hypothesis that a dimensional approach to NDs may enable us to gain a deeper understanding of individual wellbeing.

### Cognitive Complexity in Neurodiverse Diagnoses

According to a National Institute of Health (NIH) panel of experts, the cognitive domains that are most important for health and overall daily functioning, as well as academic and professional attainment, are the domains of executive function, attention, memory, language and processing speed (Weintraub et al., 2013). Differences in these cognitive domains have been shown to contribute to considerable variability in individual profiles across NDs.

Executive functions (EF) and attention are widely implicated in NDs, although findings are often inconsistent. Difficulties in inhibition, working memory, and cognitive flexibility have been linked to ASD, dyslexia, and ADHD (Howard et al., 2023; Smith-Spark et al., 2016; Kofler et al., 2010), yet not all individuals with these diagnoses show difficulties in these areas (e.g., Biederman et al., 2007; Holmes et al., 2010; Booth et al., 2010). Processing speed difficulties have also been observed across several NDs, including autism, ADHD and dyslexia (Zapparrata et al., 2023; Mostert et al., 2015; Dubois et al., 2010), and have been shown to interact with other cognitive differences in neurodivergent populations, such as working memory or attention difficulties (e.g., Kibby et al., 2019; Jacobson et al., 2011). Interestingly, evidence suggests that both slowed processing speed and EF difficulties may play a significant role in the overlap or co-occurrence of NDs, such as ADHD and dyslexia (e.g., Willcutt et al., 2010; Lonergan et al., 2019).

Moreover, differences in episodic memory (the ability to recall personal experiences) have also been observed across NDs, though findings vary. For instance, autistic individuals may struggle to recall contextual details of previous events (Bowler et al., 2011), with research suggesting that this difficulty may be related to executive function and cognitive control difficulties characteristic of ASD (Maister et al., 2013). Conversely, some individuals with dyslexia have been shown to display enhanced episodic memory abilities, potentially as a compensatory strategy (Eide & Eide, 2011). Finally, language difficulties are also common in neurodiverse populations, and vary widely across and within NDs. For instance, dyslexia is inherently a language-based condition, characterised by phonological, vocabulary and syntax difficulties (e.g., Lyon et al., 2003; Snowling et al., 2003), while ADHD has also been associated with difficulties in language comprehension (McInnes et al., 2003), despite language difficulties not being part of the diagnostic criteria for this ND (APA, 2013). Research has shown that autistic children may also experience language difficulties, with the co-occurrence of communication difficulties (e.g., apraxia of speech) also being common in autism (Vogindroukas et al., 2022). Notably, overlaps between ADHD, dyslexia and Specific Language Impairment are high, especially when conditions co-occur (Helland et al., 2016).

Importantly, these domains often interact, rather than function independently. For instance, the associations between language and cognitive development have been well established (Zauche et al., 2016), with EF differences being linked to difficulties with language processing in both ASD and ADHD (Weismer et al., 2018; Hawkins et al., 2016). These interdependencies may contribute to cascading effects in development, where early differences in one domain may affect outcomes in other domains. Together, this evidence underscores the complexity of cognitive functioning in NDs, and highlights that specific cognitive difficulties are not confined to diagnostic categories, but are transdiagnostic in nature, thus contributing to both within-diagnosis variability and between-diagnosis overlaps. This challenges the utility of studying these domains strictly in isolation or in case-control studies with narrowly defined diagnostic categories. A dimensional, transdiagnostic approach that explores interactions between these processes across a broad spectrum of individuals may be better suited to capturing the complexity of cognitive functioning in neurodivergent populations. The identification of shared and/or distinct cognitive profiles may allow for a more nuanced understanding of individual wellbeing, and potentially inform more tailored support strategies.

### Co-occurring Mental Health Conditions

Neurodiverse diagnoses not only co-occur with conditions within the same diagnostic grouping (i.e., the co-occurrence of two NDs, such as ADHD and dyslexia), but also with conditions that may not be within the same diagnostic group (England-Mason, 2020). Research suggests that NDs most often co-occur with affective and/or mental health difficulties, many of which can be categorised into ‘internalising’ and ‘externalising’ conditions. For instance, a large body of literature has demonstrated that youth with learning difficulties (e.g., dyslexia) experience elevated levels of depressive and/or anxious symptomatology (i.e., internalising problems) relative to their typically developing peers (Nelson & Harwood, 2011). Similarly, externalising difficulties (e.g., conduct problems) have been shown to be the most frequent co-occurring conditions in children with ADHD (Jensen & Steinhausen, 2015), whilst both internalising and externalising problems have been reported to be the most prevalent co-occurring conditions in autism (van Steensel et al., 2013; Gjevik et al., 2011; Underwood et al., 2019). Further, emotion regulation problems have frequently been recognised as a common difficulty across various NDs, and have been shown to be associated with both internalising and externalising conditions in neurodivergent individuals (Graziano & Garcia, 2016; Kouvava et al., 2022; Boyes et al., 2020). This section will briefly review the co-occurrence of NDs with internalising and externalising difficulties, and consider how emotion regulation may underpin complex patterns of co-occurring conditions across NDs.

#### Internalising

Internalising problems, such as anxiety and depression, involve disturbances in mood and emotional states (Zahn-Waxler et al., 2000). Research has shown that features of NDs may increase individual susceptibility to experiencing internalising difficulties (Baraskewich & McMorris, 2019). For instance, intolerance of uncertainty – a key characteristic of autism – has been suggested as a key mechanism underpinning the co-occurrence of anxiety in autistic individuals (APA, 2013; South & Rodgers, 2017; Hwang et al., 2020). Similarly, reading difficulties characteristic of dyslexia have been shown to be associated with increased levels of anxiety (Zuppardo et al., 2021), further highlighting how neurodiverse difficulties may contribute to the development internalising problems. Internalising problems may also exacerbate cognitive challenges. For example, anxiety has been shown to increase working memory problems in adolescents with ADHD (D’Agati et al., 2019), potentially leading to more complex profiles of difficulties. This may have important implications for the support of neurodivergent individuals, with research suggesting that there is currently a lack of adequate assessment and support techniques addressing more complex needs resulting from the co-occurrence of NDs and internalising problems (e.g., Rodgers & Ofield, 2018; Koyuncu et al., 2022). Further, internalising difficulties may be under-recognised in clinical settings due to overlapping symptoms with NDs (e.g., Grogan et al., 2018). For example, restlessness could reflect hyperactivity in ADHD or physiological arousal linked to anxiety (Adwas et al., 2019; Baraskewich & McMorris, 2019; Martella et al., 2020). This diagnostic ambiguity may further complicate the identification and support of NDs. Given the significant impact of internalising problems on the quality of life of neurodivergent individuals (Rogers & Ofield, 2018), a deeper understanding of the interplay between internalising and NDs is needed in order to inform more adequate assessment and support.

#### Externalising

Externalising behaviours include aggression, hyperactivity, and disruptive tendencies, as well as lack of self-control, problems with emotion regulation, and impulsivity (Hinshaw, 1987; Cai et al., 2021; Berger & Buttelmann, 2022; Martel et al., 2017). These behaviours frequently co-occur with NDs, with estimated rates suggesting that 30-50% of individuals with ADHD also meet criteria for an externalising condition, such as oppositional defiant disorder or conduct disorder (Gnanavel et al., 2019). Autism has similarly been associated with elevated levels of aggression and self-injurious behaviour (Soke et al., 2016; Kanne & Mazurek, 2011). Moreover, research suggests that developmental timing plays a role in the relationship between NDs and externalising behaviours. Longitudinal evidence suggests a bidirectional relationship, as externalising behaviours in childhood have been shown to predict later ADHD symptoms, and vice versa (Kuja-Halkola et al., 2015). Further, more complex ND profiles (e.g., co-occurring ADHD and dyslexia) have been associated with heightened externalising behaviours, such as impulsivity (Duranović et al., 2023). Similarly, some studies have reported higher levels of externalising behaviour in co-occurring ASD and ADHD groups, compared to ASD or ADHD alone (e.g., Berenguer et al., 2018; Tureck et al., 2013). This is especially important to consider given that neurodivergent individuals are more likely to present with a combination of NDs, as opposed to having a single ‘pure’ diagnosis (Bonti et al., 2024). Together, this evidence highlights the importance of considering the role of externalising in NDs across different stages of development, especially in individuals with more complex profiles of co-occurring conditions.

#### Emotion Regulation

Emotion regulation (ER) refers to the ability to monitor, evaluate and modify emotional responses and experiences (Thompson et al., 2008). Adaptive ER strategies (e.g., cognitive reappraisal) support effective functioning, while maladaptive strategies (e.g., worry, rumination) are linked to emotion dysregulation (Gross & John, 2003; Kashdan et al., 2006; Aldao et al., 2010). Research has consistently shown that neurodivergent individuals employ less-adaptive ER strategies. For example, children with ADHD show difficulties in understanding and recognising emotions, contributing to problems with social interactions (Graziano & Garcia, 2016), with some authors proposing that the emotional traits (e.g., impulsivity/irritability) resulting from poor ER in ADHD should be considered as ‘core symptoms’ of the diagnosis (Faraone et al., 2019). Similarly, autistic individuals often display maladaptive ER patterns, with research suggesting that poor ER may be a core feature of autism, and underpin some of the socioemotional and/or behavioural difficulties characteristic of this conditions (Cai et al., 2018; Mazefsky et al., 2013). These findings suggest that ER may be a shared mechanism underlying some of the functional difficulties associated with NDs.

Many studies have conceptualised ER as a transdiagnostic process due to its association with a broad range of diagnoses. For instance, Jaisle and colleagues (2023) recently provided evidence for the role of emotion dysregulation in both ASD and ADHD. They demonstrated that the hyperactivity and impulsivity traits of ADHD, and the restricted and repetitive behaviours characteristic of ASD, were related to social competence indirectly through emotion dysregulation. Moreover, ER difficulties have been associated with both internalising and externalising problems in neurodivergent and neurotypical individuals, with ER being a unique contributor to individual differences in the severity of such difficulties, above and beyond diagnostic status (Cai et al., 2021). Further, ER problems have been associated with cognitive difficulties in NDs. For instance, executive function has been suggested to predict the ability to regulate emotions (Schmeichel & Tang, 2015), with evidence demonstrating that neurodivergent children with greater cognitive control difficulties are more likely to display poorer ER, resulting in internalising/externalising problems (Tajik-Parvinchi et al., 2021). Similarly, the severity of internalising problems has been shown to be associated with more difficulties in executive function, through increased emotion dysregulation and ADHD traits (Battistutta et al., 2021). This evidence underscores the dynamic relationship between cognitive and emotional processes in neurodivergent populations, which may be important to consider in the assessment and support of NDs.

### Neurodiversity in Adults

There is currently an evident lack of research into how neurodiverse conditions present in adulthood, with the majority of research being conducted with child or adolescent populations. The lack of research into the functioning of neurodivergent adults has been observed across a wide range of NDs (including autism, dyslexia, and ADHD), and has had a substantial adverse impact on the identification of such conditions in adulthood (Lipinski et al., 2022; Sadusky et al., 2021; Goodman et al., 2024). The diagnostic assessment of many NDs is not only grounded in categorical research, but the vast majority of diagnostic criteria are also based on childhood presentations of such conditions (APA, 2013). This is an issue, since the developmental trajectories of many cognitive and mental health processes associated with NDs are dynamic, and inclined to change over time (e.g., Hens & Goidsenhoven, 2023). Thus, the diagnosis of neurodiverse conditions appears to be even more challenging in adulthood, with this difficulty being further exacerbated by the complex and heterogeneous nature of such conditions (Astle et al., 2022). Consequently, whilst research has frequently provided evidence for a lack of access to adequate support in neurodivergent children (Astle et al., 2022), the current state of literature and diagnostic procedures indicates that this may be even more difficult for neurodivergent adults. To this end, research suggests that such lack of adequate support may be even more critical for neurodivergent adults with co-occurring mental health conditions (Camm-Crosbie et al., 2018), emphasising the need for further research into both the cognitive and mental health processes of neurodivergent adults.

### The Present Study

There is an evident need for more comprehensive data-driven accounts of neurodiverse conditions that promote a shift away from strictly categorical research (Astle et al., 2022). This is especially important for gaining a deeper understanding of the complex cognitive profiles associated with neurodiverse diagnoses, and the variable interactions between cognitive and mental health processes that often have a significant impact on individual functioning and lived experience (Astle et al., 2022; Rodgers & Ofield, 2018). Given the developmental nature of neurodiverse conditions and the lack of information on how they present in adulthood (e.g., Nigg et al., 2020; Hens & Goidsenhoven, 2023; Lipinski et al., 2022), it is particularly important to collect empirical data capable of enabling more precise characterisation of cognitive and mental health functioning in neurodivergent adults.

Therefore, with the aim to build upon the emerging quantitative research framework, the present study adopted a transdiagnostic dimensional approach to characterising the complexity of the cognitive and mental health dimensions that underpin broadly sampled NDs. This study employed a combination of transdiagnostic sampling strategies, dimensional measures, and data reduction methods to investigate naturally occurring groups within the dataset through capturing distinct patterns of correlation and between-variable interactions. These techniques offered the potential to elucidate the mechanisms underlying the complexity of NDs, beyond that offered by traditional case-control designs. Ultimately, the aims of the transdiagnostic study were to address the following research questions:

1. In what ways do neurodivergent adults differ on key cognitive and mental health dimensions?
2. How can individual profiles be best represented in a multidimensional space of strengths and difficulties?
3. Can data-driven transdiagnostic clusters be used to better characterise individual functioning compared to traditional diagnostic categories? (Do data-driven clusters map directly onto traditional diagnostic categories?)
4. How can transdiagnostic dimensional research frameworks help to elucidate the mechanisms underlying the complexity of NDs (in terms of between-diagnosis overlaps, individual variation, and/or co-occurring conditions)?

## Materials and Methods

### Participants

A cohort of 175 adults (aged 18-51, M_age_ 21.7), who had been referred for assessment and/or sought help from support services for suspected or previously diagnosed neurodiverse conditions, were recruited using convenience sampling. The sample size was established based on previous research guidance suggesting *n* = 20-30 observations per expected subgroup when conducting cluster analysis, as this provides sufficient power for detecting number of clusters and individual cluster membership (Dalmaijer et al., 2022). This study aimed to explore a total of 8 core variables (5 cognitive and 3 mental health domains) on which individuals may differ; thus, a sample of 160 to 240 participants was deemed desirable. Participants were predominantly recruited from [*University name redacted for peer review*], including both staff and students (undergraduate and postgraduate). The study was advertised via a range of outlets, including online recruitment and mailouts to special interest groups. A smaller proportion of participants were recruited from external sources, including through neurodiversity charities (including Embracing Complexity, Dyspraxia Foundation, UK Adult ADHD Network, ADHD Adult UK, and All Age Autism), research databases (the Cambridge Autism Research Database), local support groups (The Recovery College), and via social media (LinkedIn, X, Instagram and Facebook). Participants were reimbursed for their time. The exclusion criteria for the study were as follows:

a. Non-native English speakers (English did not exclusively have to be their first language).
b. Persons with a history of traumatic brain injury with long-term effects on mental functioning.
c. Adults above 50 years of age.
d. Adolescents below the age of 16 who could not provide independent informed consent.
e. Adults with reduced capacity to provide informed consent on their own behalf (e.g., due to pervasive intellectual disability).
f. Individuals with uncorrected primary sensory impairments (e.g., hearing loss).

These exclusionary criteria were put in place as the abovementioned factors have the capacity to affect performance on the measures administered. The characteristics and diagnostic status of the sample are presented in Tables 1-2.

**Table 1.** Demographic characteristics of the sample (*n* = 175).

|  | n | % |
| --- | --- | --- |
| **Sex** |  |  |
| Male | 36 | 20.6 |
| Female | 139 | 79.4 |
| **Race** |  |  |
| Caucasian | 49 | 28.0 |
| Asian | 81 | 46.3 |
| African/Caribbean | 35 | 20.0 |
| Mixed Heritage | 10 | 5.7 |
| **First Language** |  |  |
| English | 143 | 81.7 |
| Other | 32 | 18.3 |
| **Current/Completed Education Level** |  |  |
| Secondary School | 2 | 1.1 |
| Sixth Form/College | 2 | 1.1 |
| Undergraduate Degree | 152 | 86.9 |
| Postgraduate Degree | 19 | 10.9 |
| **Referred for Difficulties** |  |  |
| No | 98 | 56.0 |
| Yes (Type of Difficulty): | 77 | 44.0 |
| Attention | 46 | 59.7 |
| Memory | 18 | 23.4 |
| Learning | 22 | 28.6 |
| Reading | 25 | 32.5 |
| Writing | 1 | 1.3 |
| Language/Speech | 2 | 2.6 |
| Visual Processing | 2 | 2.6 |
| Movement Co-Ordination | 3 | 3.9 |
| Anxiety/Depression/Stress | 4 | 5.2 |
| Impulsivity | 1 | 1.3 |
| Socialisation/Interaction | 1 | 1.3 |
| **Referred By** |  |  |
| Self | 36 | 46.8 |
| Parents/Family Member/Friends | 27 | 35.1 |
| School/College | 20 | 26.0 |
| GP/Therapist/Psychiatrist | 7 | 9.1 |
| Not Specified | 1 | 1.3 |
| **Referred To** |  |  |
| Education Services | 34 | 44.2 |
| Health Services | 43 | 55.8 |
| Local Support Services | 4 | 5.2 |
| Diagnostic Assessment | 2 | 2.6 |
| Private Support/Specialist | 3 | 3.9 |
| Not Specified | 1 | 1.3 |

**Note.** In the ‘Difficulties With’ subsection, the majority of participants reported multiple difficulties. Therefore, *n* does not correspond to the total number of referred individuals. The percentages in this section indicate the number of participants who reported a given difficulty, from the total number of referred participants. In the “Referred By” and “Referred To” sections, some individuals reported being referred by/to multiple sources. Therefore, *n* does not correspond to the total number of referred participants. Table 1 is continued below.

**Table 1 Continued.** Demographic characteristics of the sample (*n* = 175).
|  | *n* | % |
| --- | --- | --- |
| **Birth Complications** |  |  |
| No | 163 | 93.1 |
| Yes | 12 | 6.9 |
| **Family History of Difficulties** |  |  |
| No | 102 | 58.3 |
| Yes (Type of Difficulty): | 73 | 41.7 |
| Neurodivergent | 37 | 50.7 |
| Mental Health Condition | 39 | 53.4 |
| Learning Difficulty | 18 | 24.7 |
| Not Specified | 2 | 2.7 |
| **Difficulties Reported** |  |  |
| No | 24 | 13.7 |
| Yes (Type of Difficulty): | 151 | 86.3 |
| Communicating Needs | 47 | 31.1 |
| Understanding People's Behaviour | 32 | 21.2 |
| Functioning Independently | 21 | 13.9 |
| Self-Care | 47 | 31.1 |
| Understanding New Information | 63 | 41.7 |
| Co-ordinating Movements | 19 | 12.6 |
| Controlling Behaviour | 29 | 19.2 |
| Learning New Skills | 27 | 17.9 |
| Attention/Concentration | 16 | 10.6 |
| Organisation | 4 | 2.6 |
| Memory | 8 | 5.3 |
| Emotion Regulation | 2 | 1.3 |
| Overstimulation | 5 | 3.3 |
| Rigidity to Routine | 2 | 1.3 |
| Hyperactivity/Impulsivity | 3 | 2.0 |
| Reading | 4 | 2.6 |
| Writing | 27 | 17.9 |
| Maths | 3 | 2.0 |

**Note.** In the “Type of Difficulty” subsection of “Family History of Difficulties”, *n* does not correspond to the total number of participants with a family history of difficulties, as most participants reported a history of multiple difficulties. The percentages indicate the number of individuals who reported a given difficulty out of the total number of participants with a family history of difficulties. The final “Difficulties Reported” section refers to the general difficulties reported by participants, for which they may or may not have been referred in the past. The majority of participants reported having experienced multiple difficulties. Therefore, in the “Type of Difficulty” subsection, *n* does not correspond to the total number of participants who reported having experienced difficulties. The percentages indicate the number of individuals who reported each type of difficulty, out of the participants who reported having experienced difficulties.

**Table 2.**
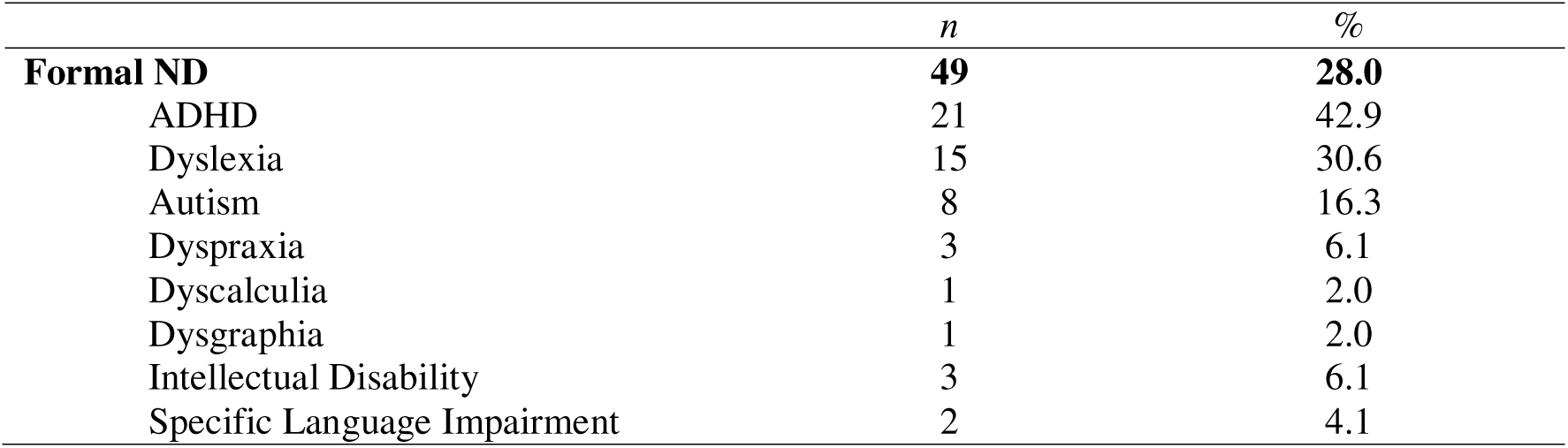
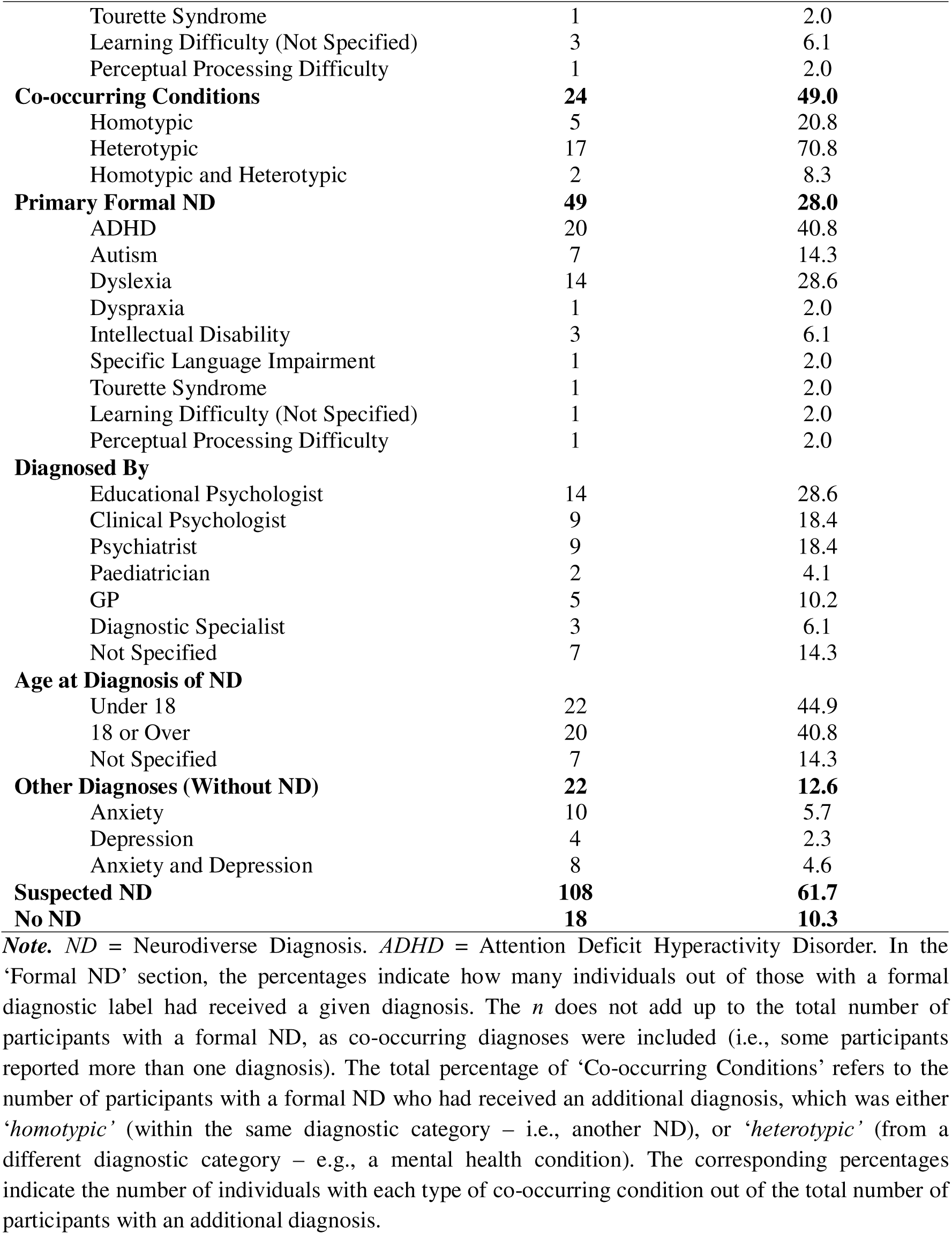
Formal diagnostic status of the sample (*n* = 175).

|  | *n* | % |
| --- | --- | --- |
| **Formal ND** | **49** | **28.0** |
| ADHD | 21 | 42.9 |
| Dyslexia | 15 | 30.6 |
| Autism | 8 | 16.3 |
| Dyspraxia | 3 | 6.1 |
| Dyscalculia | 1 | 2.0 |
| Dysgraphia | 1 | 2.0 |
| Intellectual Disability | 3 | 6.1 |
| Specific Language Impairment | 2 | 4.1 |
| Tourette Syndrome | 1 | 2.0 |
| Learning Difficulty (Not Specified) | 3 | 6.1 |
| Perceptual Processing Difficulty | 1 | 2.0 |
| **Co-occurring Conditions** | **24** | **49.0** |
| Homotypic | 5 | 20.8 |
| Heterotypic | 17 | 70.8 |
| Homotypic and Heterotypic | 2 | 8.3 |
| **Primary Formal ND** | **49** | **28.0** |
| ADHD | 20 | 40.8 |
| Autism | 7 | 14.3 |
| Dyslexia | 14 | 28.6 |
| Dyspraxia | 1 | 2.0 |
| Intellectual Disability | 3 | 6.1 |
| Specific Language Impairment | 1 | 2.0 |
| Tourette Syndrome | 1 | 2.0 |
| Learning Difficulty (Not Specified) | 1 | 2.0 |
| Perceptual Processing Difficulty | 1 | 2.0 |
| **Diagnosed By** |  |  |
| Educational Psychologist | 14 | 28.6 |
| Clinical Psychologist | 9 | 18.4 |
| Psychiatrist | 9 | 18.4 |
| Paediatrician | 2 | 4.1 |
| GP | 5 | 10.2 |
| Diagnostic Specialist | 3 | 6.1 |
| Not Specified | 7 | 14.3 |
| **Age at Diagnosis of ND** |  |  |
| Under 18 | 22 | 44.9 |
| 18 or Over | 20 | 40.8 |
| Not Specified | 7 | 14.3 |
| **Other Diagnoses (Without ND)** | **22** | **12.6** |
| Anxiety | 10 | 5.7 |
| Depression | 4 | 2.3 |
| Anxiety and Depression | 8 | 4.6 |
| **Suspected ND** | **108** | **61.7** |
| **No ND** | **18** | **10.3** |

*Note.* *ND* = Neurodiverse Diagnosis. *ADHD* = Attention Deficit Hyperactivity Disorder. In the ‘Formal ND’ section, the percentages indicate how many individuals out of those with a formal diagnostic label had received a given diagnosis. The *n* does not add up to the total number of participants with a formal ND, as co-occurring diagnoses were included (i.e., some participants reported more than one diagnosis). The total percentage of ‘Co-occurring Conditions’ refers to the number of participants with a formal ND who had received an additional diagnosis, which was either ‘*homotypic*’ (within the same diagnostic category – i.e., another ND), or ‘*heterotypic*’ (from a different diagnostic category – e.g., a mental health condition). The corresponding percentages indicate the number of individuals with each type of co-occurring condition out of the total number of participants with an additional diagnosis.

Note that participants with multiple diagnostic labels were categorised into ‘**Primary Formal ND**’ groups based on the age at which they received their diagnoses. For instance, if an individual with co-occurring conditions received a diagnosis of ADHD first, this was regarded as their ‘primary’ formal diagnosis. The corresponding percentages indicate how many individuals out of the participants with a formal ND had received a given diagnosis first. The percentages in the ‘Diagnosed By’ section indicate how many of those with a formal ND were diagnosed by each type of professional. The percentages in the ‘Other Diagnoses’ section were calculated from the number of participants out of the total sample who had received a formal diagnostic label other than ND. The ‘**Suspected ND**’ group consists of individuals who had reported one or more difficulties (see Table 1), been referred to support services and/or assessment (but never received a formal diagnosis), or are currently waiting for assessment. The ‘**No ND**’ group consists of participants who had not reported any difficulties, referrals or any ND. This group may include some individuals with a mental health condition (e.g., anxiety), who did not report any difficulties associated with NDs. The percentages in these sections indicate how many participants out of the total sample reported a suspected ND or no neurodiverse diagnosis.

### Procedure

This study received ethical approval by the [*redacted for review*] Research Ethics Committee (#HLS21066). All participants provided informed consent and completed a short demographic questionnaire online, prior to taking part in this study. All participants reconsented upon arrival on the day of their assessment to ensure that they were still happy to participate, and to confirm that participant identity was being linked with corresponding data across all measures. All participants were debriefed upon study completion, and received a brief assessment report. The cognitive and mental health batteries administered are described below.

#### Cognitive Assessment: NIH Toolbox Cognition Battery (Weintraub et al., 2013)

The National Institutes of Health (NIH) Toolbox Cognition Battery is a neuropsychological measure consisting of seven standardised assessments, tapping the overarching cognitive domains of *attention, executive function, memory, processing speed* and *language* (Weintraub et al., 2013). The battery is a computerised tool, administered to participants via an iPad, and takes approximately 36 minutes to complete. The NIH Toolbox was selected for this study due to its broad coverage of cognitive abilities, which have been shown to be relevant to a wide range of neurodiverse conditions (see Introduction). The specific cognitive functions assessed by each task are outlined below. Further, the battery has been validated for use with adults from a variety of ethnic and educational backgrounds, as well as both neurotypical and neurodivergent populations (Mungas et al., 2014; Beaumont et al., 2013; Jones et al., 2022), making it appropriate for use with more inclusive, heterogeneous samples. Additionally, the toolbox automatically generates norm-referenced continuous scores on each measure, capturing both strengths and weaknesses in cognitive functioning, making it well aligned with dimensional research frameworks. The following information has been retrieved from the NIH Toolbox Administrator’s Manual and Weintraub and colleagues (2013). *All NIH Toolbox-related materials are ©2015 Northwestern University and the National Institutes of Health.* For reference, the cognitive domains tapped by each measure are summarised in Table 3.

**Table 3.** Primary cognitive domains assessed by each cognitive task.

| Task | Domains Assessed | Description |
| --- | --- | --- |
| Flanker Inhibitory Control and Attention Test | Inhibitory Control | The ability to filter out irrelevant stimuli in the environment |
| Dimensional Change Card Sort Test | Cognitive Flexibility | The ability to efficiently switch attention in response to changing demands |
| Picture Sequence Memory Test | Episodic Memory | The ability to acquire, store and retrieve new information about personal experiences |
| List Sorting Working Memory Test | Working Memory | The ability to store information in the mind |
| Pattern Comparison Processing Speed Test | Processing Speed | The rate at which information can be received, processed and responded to |
| Picture Vocabulary Test | Vocabulary (Receptive Language) | The ability to understand the meaning of language |
| Oral Reading Recognition Test | Reading Decoding (Expressive Language) | The ability to translate written language into spoken words |

**Note.** There is substantial overlap between the cognitive domains tapped by each of the above tasks as cognitive skills can rarely be assessed in isolation due to their interdependence. For example, both the Flanker Inhibitory Control and Attention Test and the Dimensional Change Card Sort Test measure executive function as they both measure different aspects of attention. However, each of the NIH Toolbox tasks have been designed to primarily tap a specific cognitive domain (displayed above with descriptions). Therefore, each of these tasks have been retained as separate variables representing each of the cognitive processes.

#### Attention and Executive Function

The *Flanker Inhibitory Control and Attention Test* (Zelazo et al., 2014) is an adapted version of the Eriksen flanker task (Eriksen & Eriksen, 1974). The measure assesses the allocation of an individual’s limited capacities to deal with environmental stimulation, through inhibiting visual attention to irrelevant task dimensions. Thus, the task assesses inhibitory control, a component of executive function. Each trial consists of a central directional target, which is flanked by similar stimuli on each side. The participant is required to indicate the direction of the central stimulus and ignore the direction to which the arrows are pointing. During congruent trials, the flankers and target face the same direction; during incongruent trials, the target and flankers point to opposite directions. Adult performance is assessed via reaction time (RT), where scores are yielded from 0 to 10. Overall, the measure consists of 40 trials, averaging around 4 minutes to complete.

The *Dimensional Change Card Sort Test* (Zelazo et al., 2014) is a measure of set shifting (i.e., cognitive flexibility), an executive function component. The test requires the participant to match a visual stimulus to one of two choices of stimuli, according to shape or colour. Adults receive a mixed block of trials, in which colour is relevant in most trials, with occasional/unpredictable shifts to shape. The appropriate criterion word (i.e., ‘colour’ or ‘shape’) is presented on the screen, according to which participants must match the stimuli. There is a total of 40 trials which require a total of 4 minutes to complete. Performance is assessed via RT.

#### Memory

The *Picture Sequence Memory Test* (Dikmen et al., 2014) is a measure of episodic memory. The stimuli used in this measure are pictured objects/activities, which may be thematically related but have no set order. At the beginning of each trial, pictures are displayed in the centre of a computer screen and moved one at a time into a box (in a fixed order), while an audio describes the content of the picture (e.g., “Go for a hike”). This continues until the entire sequence of activities is presented on the screen. The pictures are then returned to the centre of the screen in a random display, at which point the participant is required to move them into the sequence previously demonstrated. The score is calculated from the cumulative number of adjacent picture pairs recalled correctly by the participant over three learning trials. Levels of task difficulty vary according to age group. For participants aged 9 to 60 years, 15 pictures are presented. This task takes approximately 10 minutes to complete.

The *List Sorting Working Memory Test* (Tulsky et al., 2014) is a measure of working memory. A series of stimuli are, one at a time, displayed on a computer screen both visually and orally. Participants must repeat the stimuli in size order (smallest to largest). In condition one, all stimuli presented come from one category. In the second condition, stimuli are presented from two categories. Here, the participant is required to first report all stimuli from one category, then from the other, in order of size within each. For example, if foods (category one) and animals (category two) are presented – e.g., ‘elephant’, ‘pumpkin’, ‘strawberry’ and ‘duck’ – the participant would respond with “strawberry, pumpkin, duck, elephant”. The number of items in a series increases with trials, and the test must be discontinued if the participant fails two trials of the same length. The score is derived from the total items answered correctly across trials, as scored by a trained examiner. This measure takes approximately 7 minutes to administer.

#### Processing Speed

The *Pattern Comparison Processing Speed Test* (Carlozzi et al., 2014) assesses choice reaction time (i.e., processing speed). Participants must identify whether two visual patterns are the “same” or “not the same”. The type, complexity and number of stimuli vary depending on the age spectrum from 3 to 85 years, in order to ensure adequate variability of performance. Performance is assessed via calculating the score from the total number of correct items (of a possible 130) completed in 90 seconds. This task takes approximately 3 minutes to administer.

#### Language

The *Picture Vocabulary Test* (Gershon et al., 2014) is a measure of receptive vocabulary. Single words are presented via an audio recording, and simultaneously paired with four images of different objects, actions and/or depictions of concepts. Participants are required to select the picture that most closely matches the meaning of a given word. The test items are administered in a variable-length computer-adaptive test (CAT) format, reducing the time taken to identify individual performance levels. The test uses education level to determine which items will initially be presented. This means that some participants see fewer items than others, with the specific words presented depending on the participant’s age and performance. The majority of participants will see approximately 25 items and take up to 5 minutes to complete the task. Performance is assessed based on the percentage of correct responses.

The *Oral Reading Recognition Test* (Gershon et al., 2014) assesses reading decoding skills through testing the ability to accurately pronounce single words and/or recognise letters. The test operates on a bank of English items (words/letters), controlled for orthographic typicality, frequency of word use and complexity of letter-sound relationships. The items are administered using a CAT format, which determines the items that will be presented to each participant. The participant is required to read aloud each item that is presented to them one by one on a computer screen; these responses are then entered by the examiner, who indicates whether or not the response was correct. Performance is assessed based on the number of items answered correctly, and the measure takes approximately 4 minutes to administer.

#### Scoring of the NIH Toolbox

For the purposes of this study, age-corrected standard scores were extracted from the automatically generated NIH Toolbox output. This score compares individual performance to people of the same age within the NIH Toolbox normative sample (based on the population of the United States). A score of 100 indicates that the participant performed at the national average level for their age. Scores of 15 points above or below 100 indicate that the individual’s performance was 1 standard deviation (SD) away from the average (Weintraub et al., 2013). The age-corrected standard scores were selected over uncorrected scores generated via the toolbox, as these have been shown to account for variance that is associated with non-clinical factors, providing a more accurate reflection of condition-related changes in cognitive function (Casaletto et al., 2015).

### Mental Health Assessment

The following questionnaires have been selected to form the mental health battery part of the assessment. Each measure is dimensional in nature, assessing the broad spectrum of strengths and difficulties associated with the following correlates of mental health: **internalising**, **externalising** and **emotion regulation**. All questionnaires were administered on paper and scored manually.

### The Extended Strengths and Weaknesses Assessment of Normal Behaviour (E-SWAN; Alexander et al., 2020)

The E-SWAN is an extended version of the Strengths and Weaknesses Assessment of ADHD Symptoms and Normal Behaviour (SWAN; Swanson et al., 2012). It reconceptualises DSM-5 diagnostic criteria for specific disorders as abilities ranging from high to low (i.e., strengths and weaknesses), through converting each symptom into a behaviour. The E-SWAN scales used in the current study include measures assessing ADHD, Panic Disorder, Social Anxiety and Major Depressive Disorder (MDD). The Social Anxiety and MDD scales include items assessing internalising behaviour, while the ADHD and Panic Disorder scales assess externalising symptomatology. The E-SWAN generates bidirectional distributions, reflecting the dimensional nature of psychopathology. There is a strong correspondence between the E-SWAN scores and traditional scale scores at the pathological end for each trait, with good discrimination and reliability across the full latent trait (Alexander et al., 2020). The E-SWAN was originally developed for parent-report of child behaviour, but it has been demonstrated that the framework is also valid for adult self-report (Alexander et al., 2020). Thus, the original scales have been adapted for the purposes of this study. For example, item 18 of the ADHD scale has been adapted from “Enters into conversation and games without interrupting or intruding” to “*I enter into conversation without interrupting or intruding”*. The E-SWAN is a 7-point scale. For the purposes of this study, the E-SWAN was reverse scored from 7 – ‘Far Below Average’ to 1 – ‘Far Above Average’. A mean score was calculated for each subscale. A score of 4 or above represents above average weaknesses in each area assessed, i.e., a higher score is indicative of more difficulties experienced.

### The Emotion Regulation Skills Questionnaire (ERSQ; Grant et al., 2018)

The ERSQ is an English version of a self-report measure assessing emotion regulation, originally developed and validated in German (Berking & Znoj, 2008). The questionnaire assesses 9 dimensions of emotion regulation, including awareness, sensations, clarity, understanding, acceptance, tolerance, readiness to confront, self-support and modification. Scores for each item/subscale are derived along a continuum, with the total average of items reflecting successful emotion regulation skills. The ERSQ is consistent with established measures of emotion regulation, such as the Emotion Regulation Questionnaire (Gross & John, 2003), shows good to excellent internal reliability, and has been validated for use with both typically developing and clinical adult samples (Grant et al., 2018). The ERSQ is scored on a 5-point scale ranging from 0 – ‘Not At All’ to 4 – ‘Almost Always’. The questionnaire consists of 27 items. It is scored by calculating the mean score for each subscale, with each subscale being made up of 3 items. A higher score indicates higher emotion regulation skills.

### The Strengths and Difficulties Questionnaire (SDQ; Goodman, 1997)

The SDQ is a behavioural screening questionnaire validated for adult self-report, assessing emotional symptoms, conduct problems, hyperactivity/inattention, peer relationship problems and prosocial behaviour (Goodman, 1997). The SDQ assesses both strengths and difficulties, generating a total score for general difficulties, and internalising and externalising problems, respectively. The measure has good internal consistency, test-retest stability and inter-rater agreement (Goodman, 2001), and has the capacity to predict the onset of psychopathology over time (Goodman & Goodman, 2009). The SDQ has recently been found to be equally as effective in identifying existing mental health problems in neurodivergent children without a specific diagnosis, as it has been in typically developing children and those with formal diagnoses, demonstrating its transdiagnostic utility (Bryant et al., 2020).

The SDQ consists of 25 items. It is a 3-point scale ranging from 0 – ‘Not True’ to 2 – ‘Certainly True’ (with items 7, 11, 14, 21 and 25 being reverse scored). The Emotional Difficulties, Conduct Problems, Hyperactivity, Peer Problems and Prosocial subscales each consist of 5 items and are scored out of 10 (via calculating the sum). The Externalising scale is the sum of the Conduct Problems and Hyperactivity scales, while the Internalising scale is the sum of the Emotional Difficulties and Peer Problems subscales. The Total Difficulties scale is the sum of all subscales except the Prosocial scale. The Impact score represents overall distress and impairment as a result of any difficulties experienced. A higher score on any subscale except the Prosocial scale represents a higher level of self-reported difficulties.

### Statistical Analyses

A *k*-means cluster analysis was conducted to identify data-driven groups of individuals based on shared patterns in cognitive and mental health functioning. Given the multivariate nature of the mental health dataset, a principal component analysis (PCA) was initially performed to reduce highly correlated mental health variables into fewer, orthogonal components. The identified components were then entered into the cluster analysis, along with the seven cognitive variables assessed. A one-way analysis of variance (ANOVA) was performed to explore group differences in cognition and mental health between the identified data-driven clusters, followed by Games-Howell post-hoc comparisons (Games & Howell, 1976). This test is robust to violations of normality assumptions, and thus suitable for use with mental health data generated via rating scales. All initial analyses were conducted in jamovi (Version 2.3.28; The jamovi project, 2023). Additionally, a linear discriminant analysis (LDA) was performed in SPSS (Version 29.0.2.0; IBM SPSS Statistics, 2023) to define the underlying structure of the dataset through characterising the processes (i.e., dimensions) that best differentiate between the identified clusters. Finally, the composition of the data-driven clusters was inspected to determine whether they corresponded to traditional diagnostic categories. The rationale for each data reduction technique is provided in the following sections.

#### Cluster Analysis

Cluster analysis is an unsupervised data reduction technique used to identify naturally occurring groups within a given dataset (Aldenderfer & Blashfield, 1984). It enables the discovery and characterisation of participant groups from multivariate data through capturing distinct patterns of correlation between large sets of variables that vary across groups of individuals (Kaufman & Rousseeuw, 2009). The clustering algorithm groups data into subsets based on similarity or dissimilarity, meaning the patterns of data that co-exist within a valid cluster are more similar to each other than they are to patterns belonging to different clusters within a dataset (Frades & Matthiesen, 2010). This allows for the exploration of variability across participants and measures, and the identification of variables that are the most significant differentiators between clusters.

In the process of clustering, data are not initially classified into groups based on predefined properties, which allows for any identified differences between groups to be completely data-driven (Astle et al., 2022). In this way, cluster analysis helps to elucidate how different cognitive and mental health variables may interact to form data-driven clusters, and how such processes may be associated with varying presentations of neurodiverse diagnoses (NDs).

Importantly, empirically derived groups may or may not directly map onto existing diagnostic labels. To this end, data-driven transdiagnostic clusters may be compared with traditional diagnostic categories, offering the potential to detect inaccurate diagnostic labels, challenge supposed boundaries between patterns of difficulties, or identify novel ND subtypes (Astle et al., 2022). Ultimately, clustering has the capacity to allow for a more accurate characterisation of individual strengths and weaknesses, above and beyond diagnostic status.

The application of such data-driven grouping methods has recently gained traction in the field of neurodiversity research. For example, Rivard and colleagues (2023) performed a cluster analysis to identify meaningful clinical profiles of children with suspected NDs that transcend diagnostic labels. The authors identified three clusters which significantly differed from one another, with only one of these groups aligning with a specific diagnostic category (i.e., autism). To this end, the other clusters were associated with a mix of diagnoses (e.g., ADHD, learning difficulties), and diverse difficulties (e.g., in intellectual and social functioning) that did not directly map onto specific diagnostic labels (Rivard et al., 2023). Similar results were observed by Mareva and colleagues (2024), who applied a *k*-means cluster analysis to explore data-driven profiles of executive function in a transdiagnostic sample of neurodivergent children. They identified three clusters with differing profiles, which were not directly related to diagnostic categories or to ‘core’ dimensions associated with specific diagnoses (Mareva et al., 2024). Additionally, Zdorovtsova and colleagues (2023) used cluster analysis to explore associations between inattention and hyperactivity behaviours and features of the structural brain connectome in a transdiagnostic sample of neurodivergent children. The results of this study revealed two clusters with invariant behavioural profiles (i.e., inattentive and hyperactive), which displayed significant differences in cognitive abilities (e.g., executive function). The authors suggested that inattention and hyperactivity may show overlaps across neurodiverse conditions because they can emerge through multiple trajectories of brain development (Zdorovtsova et al., 2023). Together, these findings demonstrate the insights that can be gained from data-driven grouping of transdiagnostic samples, in terms of exploring within-diagnostic heterogeneity, the mechanisms that may underpin overlaps across NDs, and individual profiles beyond diagnostic categories.

#### Principal Component Analysis (PCA)

PCA is often used in conjunction with clustering to aid in dimensionality reduction. PCA is an unsupervised data reduction technique which captures the maximum amount of variance within high-dimensional datasets through creating new uncorrelated variables (i.e., principal components), whilst minimising information loss (Jolliffe & Cadima, 2016; Greenacre et al., 2022). It is specifically useful for simplifying complex datasets in preparation for cluster analysis, as it highlights the key patterns between variables, making it easier for the algorithm to efficiently identify meaningful clusters (e.g., Ding & He, 2004). Research has shown that removing irrelevant features (i.e., those with low variance) enables the filtering of statistical ‘noise’ and results in more robust clustering solutions (Ben-Hur & Guyon, 2003). Similarly, through creating a set of uncorrelated components, PCA directly addresses the issue of multicollinearity (high correlation between several variables), which can often distort clustering algorithms and lead to unstable and/or misleading groupings (Zhang & Castelló, 2017). PCA has previously been effectively employed as a pre-processing step for cluster analysis in neurodiversity research. For instance, Zheng and colleagues (2020) performed a hierarchical cluster analysis on principal components to explore developmental and behavioural heterogeneity among autistic preschoolers. This approach enabled the identification of meaningful subgroups based on multiple measures of cognition, language, and adaptive/social skills (Zheng et al., 2020). These findings highlight the utility of PCA in extracting core dimensions from highly dimensional multivariate datasets, particularly for enhancing the reliability and interpretability of subsequent clustering solutions.

#### Linear Discriminant Analysis (LDA)

LDA is another (supervised) machine learning method used for dimensionality reduction, which explores linear combinations of features that best separate classes of data (e.g., Zhao et al., 2024). Similarly to PCA, LDA can also be utilised in conjunction with cluster analysis; for instance, it can be used to define the underlying structure of a dataset, through characterising the dimensions (i.e., discriminant functions) that best differentiate between data-driven clusters (Méndez et al., 2002). This method has recently been adopted by Zhang and colleagues (2022), who explored transdiagnostic symptom subtypes across autism and ADHD. In their study, LDA was applied to identify the cognitive and behavioural processes that best characterised data-driven clusters, providing evidence for precise phenotyping of the two NDs (Zhang et al., 2022). These findings demonstrate the value of employing LDA in conjunction with cluster analysis for investigating heterogeneity in neurodivergent populations, and for providing a more nuanced characterisation of individual profiles.

#### Addressing Research Aims

Overall, the combination of PCA, cluster analysis, and LDA was well suited for addressing the aims of this study. First, PCA was used as a pre-processing step to simplify the high-dimensional dataset, enhancing the stability and interpretability of subsequent analyses. Cluster analysis was then employed to identify naturally occurring groups, enabling a data-driven examination of whether distinct cognitive and/or mental health profiles exist among a mixed sample of neurodivergent adults, beyond the boundaries of traditional diagnostic labels. This addressed the study’s aim to explore whether individual functioning can be better characterised by data-driven clusters rather than diagnostic categories. Finally, LDA was used to interpret the resulting clusters by identifying the combinations of variables that best distinguish between them. This allowed for a deeper understanding of the strengths and difficulties along key cognitive and mental health dimensions that define each subgroup. Together, these analyses highlighted the cognitive and/or mental health mechanisms that may underpin the complexity associated with NDs (in terms of heterogeneity, overlaps, and/or co-occurrence). For instance, if a cluster predominantly composed of individuals with diagnoses of ADHD and dyslexia was characterised by a particular pattern of cognitive and/or mental health strengths and difficulties, this could point to shared underlying processes that contribute to the co-occurrence of these conditions, or reveal overlapping areas of functioning across the two ND profiles. Similarly, if individuals with the same diagnostic label were distributed across multiple data-driven clusters – each defined by a distinct profile of cognitive and/or mental health functioning – this may indicate the presence of underlying mechanisms that contribute to individual variability within diagnostic categories.

Given the exploratory and data-driven nature of cluster analysis, there were no *a priori* hypotheses regarding the number or structure of potential clusters. The main aim of this analysis was to uncover naturally occurring groups within the transdiagnostic dataset, without imposing predefined assumptions. Nonetheless, based on previous research, it was predicted that the data-driven groups would not reliably map onto the primary diagnostic labels present within the dataset.

## Results

### Principal Component Analysis (PCA)

A principal component analysis was performed on the 19 mental health variables assessed, with the aim to reduce multivariate data to a set of dimensions, which together explain the largest amount of variance within the self-reported mental health measures. The mental health variables entered into the PCA and correlations between all variables (including cognitive tasks) are presented in Table 4. Prior to the analysis, the Strengths and Difficulties Questionnaire (SDQ) and the Extended Strengths and Weaknesses Assessment of Normal Behaviour (E-SWAN) variables were rescaled such that low scores represented more difficulties (or less adaptive functioning) across all measures. Any mental health composite scores (i.e., the total score of the ERSQ, and the Total Difficulties, Externalising and Internalising subscales of the SDQ) were excluded from the analysis due to the inherent correlations between individual measures and their respective composite variables. Data were standardised across measures via converting all values to *z*-scores (with reference to the sample).

**Table 4.**
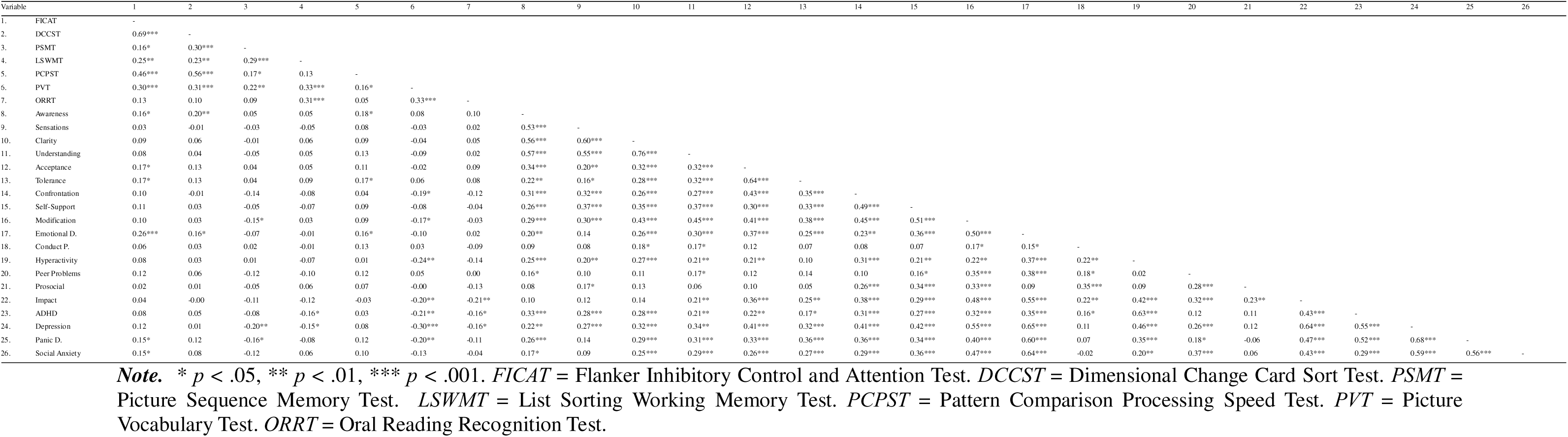
Pearson’s *r* correlation matrix for all cognitive and mental health variables (n = 175).

Bartlett’s Test of Sphericity was conducted to assess whether the correlation matrix of the mental health variables was significantly different from an identity matrix (i.e., whether the correlation coefficients between the variables were different from zero). The results of the test were significant (*x*²[171, *N* = 175] = 1563, *p* < .001), indicating the variables significantly covaried and were therefore suitable for PCA. The PCA was conducted with no statistical rotation to maximise the amount of variance explained by each component. A minimum factor loading of 0.5 was applied to highlight variables with only moderate or strong correlations to a given component. The optimal number of components retained from the analysis was determined using parallel analysis, which suggested a 4-component solution. This was validated by the finding that the first four observed eigenvalues exceeded those generated from randomly simulated data, indicating that these components explained more variance than would be expected by chance. The variable loadings on each component are presented in Table 5. The threshold of cumulative variance explained was set to 60%, as increasing this threshold resulted in the inclusion of components with factor loadings below 0.5 (i.e., no more added value). The percentage of variance explained by each principal component is presented in Table 6. Together, the first four components explained 61.1% of the total variance, with PC1 and PC2 accounting for majority of the variance in the dataset (45.7%).

**Table 5.** Loadings of mental health variables onto principal components.

| Measure | Component |  |  |  |
| --- | --- | --- | --- | --- |
|  | 1 | 2 | 3 | 4 |
| **ERSQ** |  |  |  |  |
| Awareness | **0.528** | **0.550** | -0.156 | 0.098 |
| Sensations | 0.498 | **0.611** | -0.016 | 0.148 |
| Clarity | **0.613** | **0.582** | -0.096 | 0.052 |
| Understanding | **0.627** | **0.541** | -0.113 | -0.061 |
| Acceptance | **0.593** | 0.061 | -0.077 | -0.348 |
| Tolerance | **0.520** | 0.060 | -0.102 | -0.494 |
| Confrontation | **0.604** | 0.071 | 0.083 | -0.075 |
| Self-Support | **0.624** | 0.126 | 0.182 | -0.159 |
| Modification | **0.740** | -0.007 | 0.237 | -0.203 |
| **SDQ** |  |  |  |  |
| Emotional Difficulties | **0.702** | -0.391 | -0.048 | -0.060 |
| Conduct Problems | 0.247 | 0.046 | **0.535** | 0.356 |
| Hyperactivity | **0.522** | -0.133 | -0.154 | **0.633** |
| Peer Problems | 0.378 | -0.192 | 0.441 | -0.195 |
| Prosocial Skills | 0.279 | 0.071 | **0.806** | 0.097 |
| Impact | **0.653** | -0.428 | 0.142 | 0.120 |
| **E-SWAN** |  |  |  |  |
| ADHD | **0.606** | -0.151 | -0.183 | **0.538** |
| Depression | **0.796** | -0.331 | -0.137 | 0.070 |
| Panic Disorder | **0.694** | -0.321 | -0.340 | 0.002 |
| Social Anxiety | **0.633** | -0.368 | -0.105 | -0.243 |

*Note.* *ERSQ* = Emotion Regulation Skills Questionnaire. *SDQ* = Strengths and Difficulties Questionnaire. *E-SWAN* = Extended Strengths and Weaknesses Assessment of Normal Behaviour. *ADHD* = Attention Deficit Hyperactivity Disorder. Strong correlations (above 0.5) are highlighted in bold. These correlations were used to define each component. Component scores were derived from the full set of loadings, with variables contributing proportionally based on the strength of their loadings.

**Table 6.** Percentage of variance explained by each principal component.

| Component | SS Loadings | % of Variance | Cumulative % |
| --- | --- | --- | --- |
| 1 | 6.58 | 34.61 | 34.6 |
| 2 | 2.10 | 11.07 | 45.7 |
| 3 | 1.52 | 7.98 | 53.7 |
| 4 | 1.42 | 7.49 | 61.1 |

*Note.* *SS* = Sum of Squares. The *SS* loading is the sum of squared loadings for all variables contributing to a given component. This indicates how much of the variance in the original dataset is accounted for by that component, based on the contributions of each variable. An *SS* loading close to 1 indicates that the component explains a larger amount of variance in the data.

The component loadings indicated that PC1 was primarily associated with variables relating to emotion regulation difficulties, hyperactivity, impact of difficulties and specific mental health conditions, while PC2 displayed strong correlations with variables relating to emotion regulation skills. Additionally, PC3 was strongly associated with variables relating to antisocial behaviour (‘Conduct Problems’ and ‘Prosocial Skills’), while PC4 was primarily related to ADHD and hyperactivity. It is important to note that there was an overlap in the loadings of several variables. For instance, ‘Awareness’, ‘Clarity’ and ‘Understanding’ were strongly correlated with PC1 and PC2, with ‘Awareness’ being more strongly related to PC2, and ‘Clarity’ and ‘Understanding’ having a stronger relationship with PC1. Further, ‘Hyperactivity’ and ‘ADHD’ were associated with both PC1 and PC4, with ‘Hyperactivity’ being more strongly correlated with PC4, and ‘ADHD’ showing a stronger association with PC1.

Since the variables ‘Modification’, ‘Emotional Difficulties’ and ‘Depression’ had the highest loadings onto PC1, this component was defined as the *‘Emotional Wellbeing’* dimension. Further, ‘Sensations’ and ‘Clarity’ had the highest loadings onto PC2; this component was described as the ‘*Emotional Awareness’* dimension. Additionally, ‘Prosocial Skills’ had the highest loading on PC3; this component was defined as the ‘*Social Skills’* dimension. Finally, ‘Hyperactivity’ had the highest loading onto PC4; this component was labelled as the ‘*Behavioural Regulation’* dimension.

### Cluster Analysis

A *k*-means cluster analysis was conducted to investigate the structure of the dataset through identifying distinct data-driven groups based on patterns of strengths and weaknesses in cognitive and mental health functioning. The four identified mental health components (Emotional Wellbeing, Emotional Awareness, Social Skills and Behavioural Regulation) were entered into the analysis (representing individual scores), along with seven cognitive variables, each represented by the NIH Toolbox task administered: 1) Flanker Inhibitory Control and Attention Test (FICAT), 2) Dimensional Change Card Sort Test (DCCST), 3) Picture Sequence Memory Test (PSMT), 4) List Sorting Working Memory Test (LSWMT), 5) Pattern Comparison Processing Speed Test (PCPST), 6) Picture Vocabulary Test (PVT), and 7) Oral Reading Recognition Test (ORRT). All data were standardised (i.e., converted to *z*-scores, referenced to the sample) when entered into the cluster analysis.

A three-cluster structure was suggested by the gap statistic (Tibshirani et al., 2001) and elbow method (Ketchen & Shook, 1996) as the optimal solution. The clusters consisted of 50, 73 and 52 participants, respectively (i.e., all individuals were placed in only one cluster). Table 7 shows that Cluster 2 captured the majority of individuals with a formal neurodiverse diagnosis (ND; 44.9% of all individuals with a formal diagnosis), followed by Cluster 3 (36.7%) and Cluster 1 (18.4%). Individuals with a suspected (but unconfirmed) ND were also most often members of Cluster 2 (40.7% of all individuals with a suspected ND), followed by Cluster 1 (31.5%), and Cluster 3 (27.8%). Finally, individuals with no ND (i.e., no formal/suspected diagnosis) were equally represented in Cluster 1 and Cluster 2 (38.9% of all individuals with no ND, respectively), followed by Cluster 3 (22.2%). Further, Cluster 3 captured 65.0% of all participants with a primary diagnosis of ADHD, while 71.4% of all autistic individuals were members of Cluster 2. Interestingly, individuals with a primary diagnosis of dyslexia were almost evenly distributed across the three clusters (35.7% in Clusters 1 and 2, 28.6% in Cluster 3). The distributions of other, less prevalent formal diagnostic labels are presented in Table 7.

**Table 7.** Frequency of diagnostic groups and types of primary diagnoses within individual clusters derived from cognitive data and mental health principal components (*n* = 175).

| | Cluster 1<br>($n = 50$) | Cluster 2<br>($n = 73$) | Cluster 3<br>($n = 52$) |
| --- | --- | --- | --- |
| Group | $n$ (%) | $n$ (%) | $n$ (%) |
| **Diagnostic Group** |  |  |  |
| ND | 9 (18.0) | 22 (30.1) | 18 (34.6) |
| Suspected ND | 34 (68.0) | 44 (60.3) | 30 (57.7) |
| No ND | 7 (14.0) | 7 (9.6) | 4 (7.7) |
| **Type of Diagnosis** |  |  |  |
| *N* | 9 (100) | 22 (100) | 18 (100) |
| ADHD | 1 (11.1) | 6 (27.3) | 13 (72.2) |
| Autism | 1 (11.1) | 5 (22.7) | 1 (5.6) |
| Dyslexia | 5 (55.6) | 5 (22.7) | 4 (22.2) |
| Dyspraxia | - | 1 (4.5) | - |
| ID | 1 (11.1) | 2 (9.1) | - |
| SLI | - | 1 (4.5) | - |
| TS | - | 1 (4.5) | - |
| PPD | 1 (11.1) | - | - |
| SLD | - | 1 (4.5) | - |

**Note.** *ND* = Neurodiverse Diagnosis. *ADHD* = Attention Deficit Hyperactivity Disorder. *ID* = Intellectual Disability. *SLI* = Specific Language Impairment. *TS* = Tourette Syndrome. *PPD* = Perceptual Processing Difficulty. *SLD* = Specific Learning Difficulty (but not specified). The frequency of primary diagnosis type was calculated for individual clusters from the total number of participants with a formal ND found within each cluster.

The descriptive statistics across the cognitive and mental health variables are presented in Table 8. A one-way analysis of variance (ANOVA) was conducted to explore group differences between the three clusters on these variables. The results revealed significant group differences on the Emotional Wellbeing, Emotional Awareness and Behavioural Regulation dimensions (see Table 8). Note that for the following results, a higher score on any variable indicates more adaptive functioning (fewer difficulties).

**Table 8.**
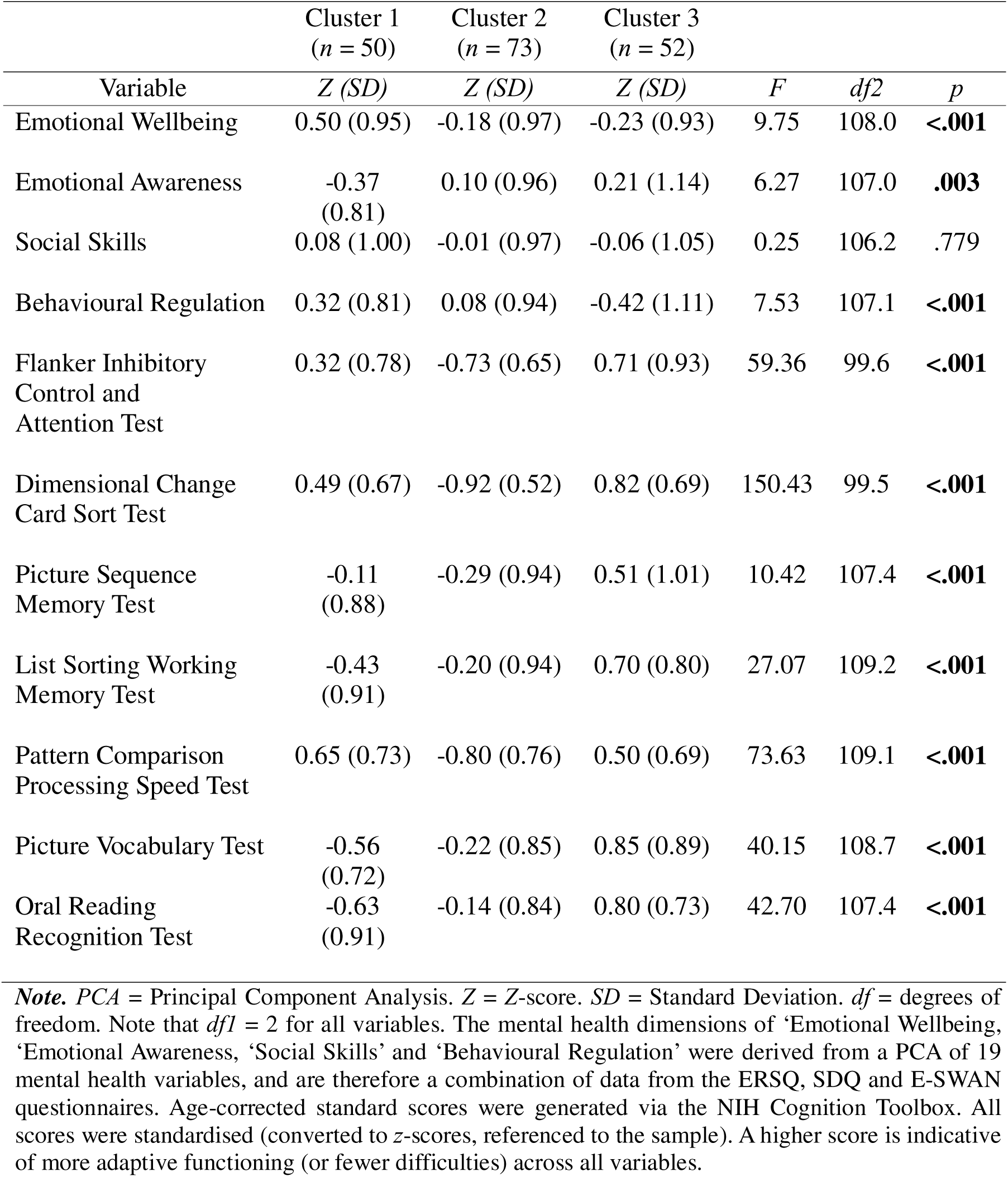
Mean *z*-scores and one-way ANOVA results for group differences across cognitive tasks and mental health dimensions for clusters derived from cognitive variables and mental health components (*n* = 175).

Post-hoc comparisons using the Games-Howell test indicated that Cluster 1 reported significantly higher Emotional Wellbeing than Cluster 2 (*t*(106) = 3.90, *p* < .001, *d* = 0.71) and Cluster 3 (*t*(99.6) = 3.90, *p* < .001, *d* = 0.78). Additionally, Cluster 1 displayed significantly lower Emotional Awareness scores than Cluster 2 (*t*(116) = -2.98, *p* = .010, *d* = - 0.52) and Cluster 3 (*t*(91.9) = -3.00, *p* = .010, *d* = -0.58). For Behavioural Regulation, Cluster 1 reported significantly fewer problems than Cluster 3 (*t*(93.6) = 3.89, *p* < .001, *d* = 0.76), and Cluster 2 also reported significantly fewer problems than Cluster 3 (*t*(98.8) = 2.67, *p* = .024, *d* = 0.49).

A one-way ANOVA also revealed significant group differences on all of the cognitive variables. Post-hoc analyses using the Games-Howell test indicated that Cluster 1 scored significantly higher than Cluster 2 on the FICAT (*t*(93) = 7.84, *p* < .001, *d* = 1.49), while Cluster 2 also displayed significantly lower scores than Cluster 3 on the same task (*t*(85.5) = - 9.57, *p* < .001, *d* = -1.85). Significant differences were also observed on the DCCST with Cluster 1 scoring significantly higher than Cluster 2 (*t*(88.1) = 12.5, *p* < .001, *d* = 2.41) and significantly lower than Cluster 3 (*t*(100) = -2.44, *p* = .043, *d* = -0.49). Cluster 2 also scored significantly lower than Cluster 3 on this task (*t*(90.6) = -15.26, *p* < .001, *d* = -2.92). For the PSMT, both Cluster 1 (*t*(99.1) = -3.34, *p* = .003, *d* = -0.65) and Cluster 2 (*t*(104.9) = -4.50, *p* < .001, *d* = -0.83) scored significantly lower than Cluster 3. Post-hoc comparisons indicated similar results on the LSWMT, with both Cluster 1 (*t*(97.4) = -6.72, *p* < .001, *d* = -1.32) and Cluster 2 (*t*(118.9) = -5.83, *p* < .001, *d* = -1.02) displaying significantly lower scores than Cluster 3. Significant differences were also found on the PCPST, with Cluster 1 scoring significantly higher than Cluster 2 (*t*(109) = 10.7, *p* < .001, *d* = 1.94), and Cluster 2 scoring significantly lower than Cluster 3 (*t*(116) = -9.96, *p* < .001, *d* = -1.78). Additionally, Cluster 1 (*t*(96.8) = -8.80, *p* < .001, *d* = -1.74) and Cluster 2 (*t*(106.2) = -6.77, *p* < .001, *d* = -1.23) scored significantly lower than Cluster 3 on the PVT. Finally, Cluster 1 scored significantly lower than Cluster 2 (*t*(99.9) = -2.97, *p* = .010, *d* = -0.56) and Cluster 3 (*t*(93.9) = -8.71, *p* < .001, *d* = -1.74) on the ORRT. Cluster 2 also scored significantly lower than Cluster 3 on this task (*t*(118.2) = -6.70, *p* < .001, *d* = -1.18).

#### Summary of Cluster Characteristics

Based on the above results, the three clusters can be described as below, based on pairwise comparisons with each other. The differences in cluster profiles across the cognitive and mental health variables are visually presented in Figure 1. For reference, the primary cognitive domains assessed by each cognitive task are presented in Table 3.

**Figure 1.**
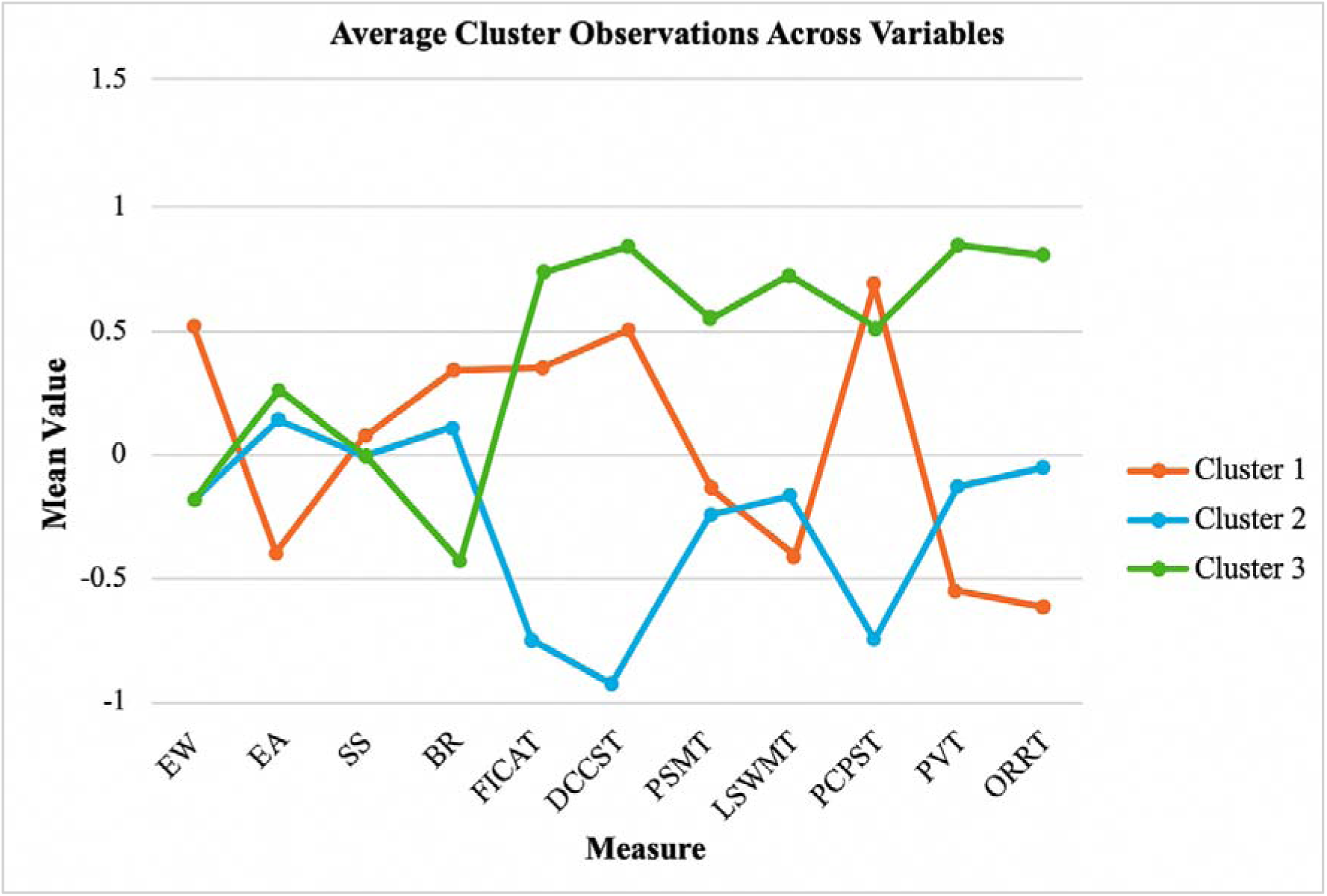
Average observations across cognitive and mental health variables for the three-cluster solution. ***Note***. *EW* = Emotional Wellbeing. *EA* = Emotional Awareness. *SS* = Social Skills. *BR* = Behavioural Regulation. *FICAT* = Flanker Inhibitory Control and Attention Test. *DCCST* = Dimensional Change Cart Sort Test. *PSMT* = Picture Sequence Memory Test. *LSWMT* = List Sorting Working Memory Test. *PCPST* = Pattern Comparison Processing Speed Test. *PVT* = Picture Vocabulary Test. *ORRT* = Oral Reading Recognition Test.

**Cluster 1**

*Mental Health:* Higher emotional wellbeing, lower emotional awareness, higher behavioural regulation.

*Cognition:* Moderate inhibitory control, moderate cognitive flexibility, weak episodic memory, weaker working memory, stronger processing speed, weaker vocabulary, very weak reading skills.

**Cluster 2**

*Mental Health:* Lower emotional wellbeing, higher emotional awareness, moderate behavioural regulation.

*Cognition:* Weaker inhibitory control, weaker cognitive flexibility, weaker episodic memory, weaker working memory, weaker processing speed, weaker vocabulary, weaker reading skills.

**Cluster 3**

*Mental Health*: Lower emotional wellbeing, higher emotional awareness, lower behavioural regulation.

*Cognition:* Stronger inhibitory control, stronger cognitive flexibility, stronger episodic memory, stronger working memory, moderate processing speed, stronger vocabulary, stronger reading skills.

### Linear Discriminant Analysis (LDA)

A linear discriminant analysis (LDA) was conducted on the three clusters derived from the four mental health components and seven cognitive variables. This was performed with the aim to characterise the discriminant functions (i.e., dimensions) that best statistically differentiate between the data-driven clusters, and therefore help to define the underlying structure of the dataset. Since LDA is a parametric method and assumes that data are normally distributed, Box’s M test was conducted to evaluate homogeneity of variance-covariance across clusters, given the heterogeneous nature of the sample. The result was not significant (*p* = .547), confirming that the assumption of equal variance-covariance was not violated.

The LDA revealed that the dataset was best defined by two discriminant functions, suggesting that the clusters were best represented in a two-dimensional space. Combined, the first two discriminant functions significantly distinguished between the three clusters (*x*²(22) = 358.48, *p* < .001), with a Wilk’s Lambda of .117. Additionally, the second discriminant function alone significantly contributed to the separation of the clusters (*x*²(10) = 138.85, *p* < .001), with a Wilk’s Lambda of .435. Function 1 explained 67.8% of the variance, while Function 2 explained the remaining 32.3% of the variance in the dataset.

The structure matrix (Table 9) revealed that the DCCST showed the strongest correlation with Function 1 (*r* = .77), followed by the PCPST (*r* = .54) and the FICAT (*r* = .50). Additionally, the canonical coefficients table (Table 10) indicated that the DCCST (β = .70, *p* < .001) and PCPST (β = .49, *p* < .001) had the largest weighting on Function 1, suggesting they were the most influential predictors in significantly differentiating between the clusters. The FICAT had a relatively low weighting on Function 1 (β = .19, *p* < .001), suggesting that although significant, it was not a strong contributor to the final classification among the other variables. Therefore, the first discriminant function (i.e., *Dimension 1*) could be defined by a combination of *cognitive flexibility* and *processing speed* abilities (see Table 3 for cognitive domains tapped by each task).

**Table 9.** Pooled within-groups correlations (structure matrix) between variables and canonical discriminant functions.

| Variable | Function 1 | Function 2 |
| --- | --- | --- |
| Dimensional Change Card Sort Test | .773* | -.085 |
| Pattern Comparison Processing Speed Test | .541* | -.260 |
| Flanker Inhibitory Control and Attention Test | .495* | -.001 |
| Picture Sequence Memory Test | .189* | .164 |
| Oral Reading Recognition Test | .181 | .534* |
| Picture Vocabulary Test | .229 | .513* |
| List Sorting Working Memory Test | .186 | .381* |
| Emotional Difficulties | .055 | -.285* |
| Hyperactive Behaviour | -.085 | -.240* |
| Emotional Clarity | -.020 | .216* |
| Social Skills | -.003 | -.048* |

**Note.** \* Largest absolute correlation between each variable and any discriminant function. The variables are ordered by absolute size of correlation within function.

**Table 10.** Standardised canonical discriminant function coefficients.

| Variable | Function 1 | Function 2 |
| --- | --- | --- |
| Emotional Difficulties | -.018 | -.328 |
| Emotional Clarity | -.195 | .278 |
| Social Skills | .064 | -.099 |
| Hyperactive Behaviour | -.076 | -.301 |
| Flanker Inhibitory Control and Attention Test | .185 | .068 |
| Dimensional Change Card Sort Test | .702 | -.153 |
| Picture Sequence Memory Test | .133 | .135 |
| List Sorting Working Memory Test | .050 | .321 |
| Pattern Comparison Processing Speed Test | .489 | -.377 |
| Picture Vocabulary Test | .009 | .491 |
| Oral Reading Recognition Test | .306 | .490 |

**Note.** Standardised canonical coefficients represent the relative importance of each variable in distinguishing between groups, with higher absolute values indicating stronger contributions to the discriminant functions.

Further, the structure matrix revealed that the ORRT (*r* = .53), PVT (*r* = .51) and LSWMT (*r* = .38) displayed the strongest correlations with Function 2, with Emotional Wellbeing (*r* = - .29) and Behavioural Regulation (*r* = -.24) also showing notable associations. The canonical coefficients table indicated that the ORRT (β = .49, *p* < .001) and the PVT (β = .49, *p* < .001) had the largest weighting on Function 2, followed by Emotional Wellbeing (β = -.33, *p* < .001), LSWMT (β = .32, *p* < .001) and Behavioural Regulation (β = -.30, *p* < .001). These results suggest that *language* and *working memory* abilities were the primary contributors in significantly differentiating between clusters along *Dimension 2*, with *Emotional Wellbeing* and *Behavioural Regulation* playing supplementary roles.

The classification table indicated an overall original accuracy rate of 97.1%, with 98.0% of Cluster 1 correctly classified, 95.9% of Cluster 2 correctly classified, and 98.1% of Cluster 3 correctly classified. The overall cross-validated accuracy rate was 96.0%, suggesting that the model also performed well on new, unseen data. These results indicate that the identified clusters were internally coherent and well-separated on the selected variables. Figure 2 visualises the extent to which the discriminant functions separated the data points into the three data-driven clusters.

**Figure 2.**
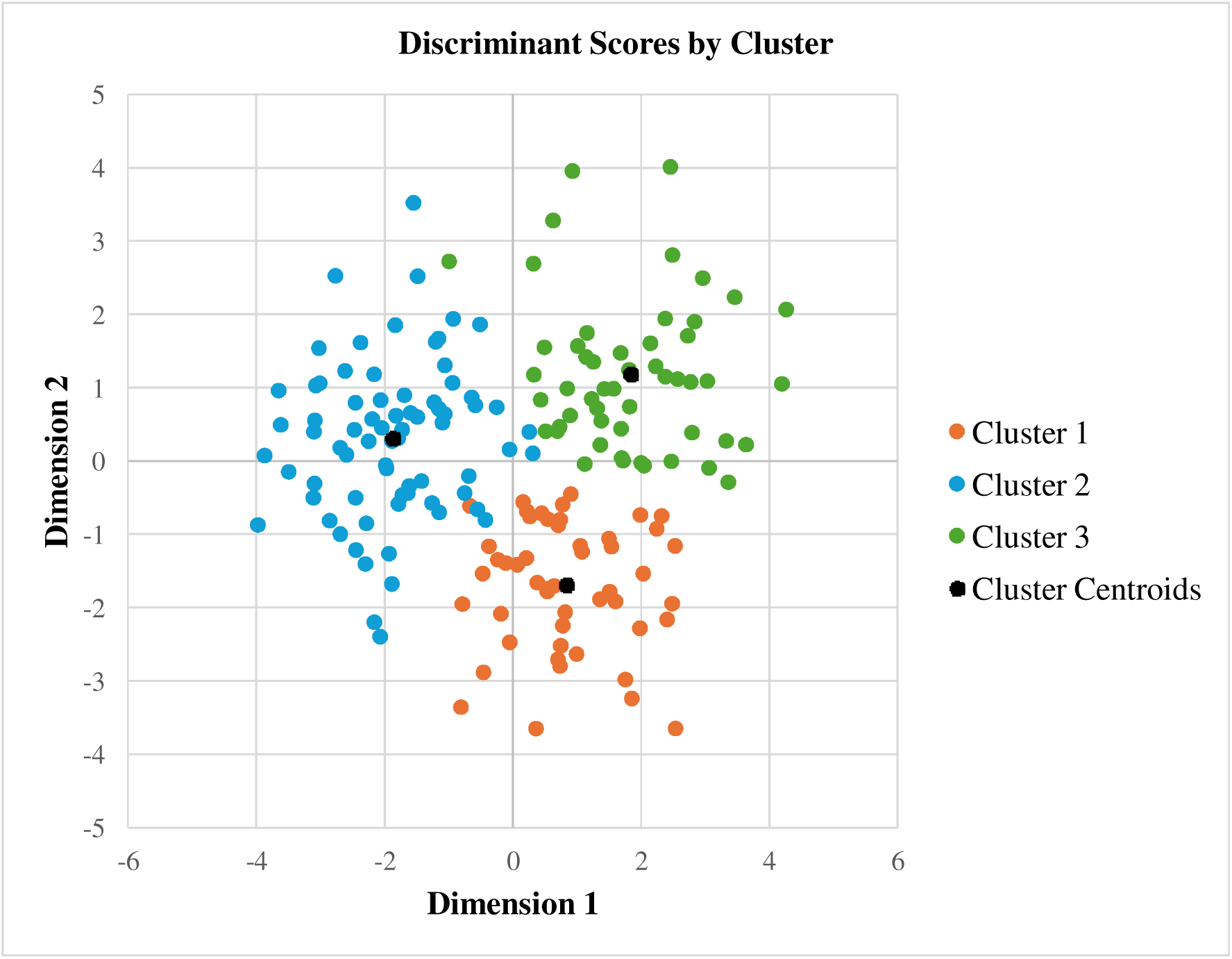
A scatterplot of discriminant function scores by data-driven cluster. ***Note.*** The axes represent discriminant scores, derived from the linear combinations of input variables that best separate the clusters. Dimension 1 = cognitive flexibility and processing speed. Dimension 2 = primarily language and working memory (along with emotional wellbeing and behavioural regulation).

Examination of the cluster centroids on each discriminant function revealed distinct profiles across the three clusters. Dimension 1 best differentiated between Cluster 2 (weaker cognitive flexibility and processing speed abilities) from Cluster 3 (stronger cognitive flexibility and processing speed), with Cluster 1 falling in the middle. It should be noted that the Emotional Wellbeing and Behavioural Regulation were negatively correlated with Dimension 2. Thus, clusters scoring high on this dimension should be interpreted as having stronger language and working memory skills, but more emotional and/or behavioural difficulties, whilst clusters scoring low on this dimension should be interpreted as demonstrating weaker language and working memory skills, but stronger emotional and/or behavioural functioning. To this end, Dimension 2 primarily separated Cluster 1 from Cluster 3, whilst Cluster 2 displayed moderate discriminant scores on this function. Cluster 1 showed weaker language and working memory abilities (with higher emotional wellbeing and behavioural regulation), whilst Cluster 3 displayed stronger language and working memory abilities (with more emotional and behavioural difficulties). Note that the mental health variables should be considered as supplementary, with Dimension 2 being primarily defined by language and working memory.

## Discussion

This study aimed to adopt a data-driven approach to grouping a transdiagnostic sample of neurodivergent adults based on patterns of strengths and difficulties across cognitive and mental health dimensions. The results revealed three data-driven groups with distinct cognitive and mental health profiles, which did not coherently map onto traditional diagnostic categories. The transdiagnostic groups were best distinguished by two core dimensions. Dimension 1 was related to cognitive flexibility and processing speed abilities. Dimension 2 was primarily defined by language and working memory skills, with emotional wellbeing and behavioural regulation also demonstrating considerable influence along this dimension. Together, these results suggest that the cognitive and mental health profiles of neurodivergent adults exist independently of diagnostic categories, and demonstrate the value of adopting transdiagnostic approaches to meaningfully characterising individual wellbeing beyond purely diagnostic frameworks.

### Individual Cluster Interpretations

Cluster 1 could be interpreted as the ‘specific learning difficulties’ (SLD) group. Although this cluster comprised the lowest number of participants with a formal neurodiverse diagnosis (ND), the most common diagnostic label found within this group was dyslexia. The cluster was primarily characterised by weak language (vocabulary, reading decoding) and working memory abilities. This profile aligns closely with the cognitive presentations of dyslexia, which has been strongly associated with difficulties in working memory, reading decoding and vocabulary (Smith-Spark & Fisk, 2007; Snowling et al., 2019; van Viersen et al., 2017). Interestingly, this group also displayed relatively moderate inhibitory control, cognitive flexibility, and intact processing speed when compared with the other clusters. This suggests that the data-driven group was characterised by specific cognitive weaknesses, rather than a ‘global’ cognitive difficulty profile. Such results are not entirely consistent with the broader literature, since many studies have provided evidence of executive function difficulties in SLD populations. For instance, cognitive flexibility and inhibition have been highlighted as specific areas of weakness in both dyslexia and dyscalculia (e.g., Barbosa et al., 2019; Zhang et al., 2025), whilst slowed processing speed has been established as a cognitive marker of dyslexia (e.g., Carroll et al., 2025). The present results could therefore underscore the heterogeneous nature of SLDs, through providing evidence of a unique cognitive profile which partially aligns with previous findings on dyslexia, whilst highlighting the variability in executive function and processing speed across individuals with SLDs. Additionally, it should be noted that Cluster 1 also captured many individuals without a formal diagnostic label, suggesting that members of this group with suspected NDs may have presented with difficulties associated with dyslexia (or other SLDs). This highlights the potential of data-driven approaches in identifying meaningful cognitive profiles in the absence of formal diagnostic labels, capturing individuals whose learning difficulties may otherwise go unrecognised in categorical research.

Further, Cluster 1 did not demonstrate a marked mental health burden. Given that this cluster captured the lowest number of individuals with formal NDs, and did not present with a broader cognitive difficulty profile, the relatively high emotional wellbeing and behavioural regulation (relative to the other clusters) would be expected, since many NDs have been associated with co-occurring mental health problems (e.g., van Steensel et al., 2013, Kouvava et al., 2022; Jensen & Steinhausen, 2015). On the other hand, the results are not in line with research evidence suggesting that individuals with dyslexia are likely to experience elevated levels of internalising problems, such as anxiety or depression (Nelson & Harwood, 2011). Thus, it is possible that Cluster 1 may represent a subset of neurodivergent adults who, despite specific learning-related difficulties, are able to maintain relatively good emotional wellbeing and behavioural regulation. This could potentially reflect successful adaptation or compensatory strategies developed over time. In fact, research has suggested that neurodivergent adults may demonstrate an improvement in emotional wellbeing across development (Woodman et al., 2016), with evidence of effective strategy development to compensate for their difficulties (e.g., Carawan et al., 2016). These findings emphasise heterogeneity within neurodivergent populations, specifically how profiles may vary or diverge in adulthood, further demonstrating the benefits of applying data-driven approaches to capture such nuanced variation across development.

Cluster 2 could be interpreted as the ‘higher need’ or ‘more severe’ group. This cluster captured the highest number of individuals with a formal ND, and was the most heterogeneous in terms of primary diagnostic labels, capturing all but one formal ND within the dataset (with ADHD, autism, and dyslexia being the most common). This group’s profile was best characterised by cognitive flexibility and processing speed weaknesses, but also displayed widespread difficulties across all cognitive domains (relative to the other two clusters). These results are in line with previous literature suggesting that the co-occurrence of conditions often results in more severe difficulties and/or more complex cognitive profiles (Mayes et al., 2000; Wang & Chung, 2018), and also aligned with research suggesting broad cognitive weaknesses across various NDs (e.g., Fuermaier et al., 2015; Howard et al., 2023; Smith-Spark et al., 2016).

Given the diversity of this data-driven group, it is possible that the combination of cognitive flexibility and processing speed difficulties may be a potential mechanism underpinning the co-occurrence of NDs. To this end, difficulties with cognitive flexibility have frequently been associated with a wide range of NDs, including autism, ADHD, and dyslexia (e.g., Lage et al., 2024; Roshani et al., 2020; Cartwright et al., 2017), and have been proposed to be a shared weakness in co-occurring NDs, such as ADHD and dyslexia (e.g., Lonergan et al., 2019). Similarly, research has shown evidence of shared processing speed weaknesses in ADHD and dyslexia (McGrath et al., 2011). It has also been suggested that individual differences in processing speed are influenced by genes that increase the risk for dyslexia, ADHD and their co-occurrence (Willcutt et al., 2010). The composition and cognitive profile of this cluster therefore provide further evidence of potential overlap between several NDs, and highlight mechanisms that may contribute to the emergence of co-occurring conditions. Such findings further demonstrate the value of data-driven approaches in capturing heterogeneity and complexity of profiles that transcend diagnostic boundaries, and provide support for a transdiagnostic, rather than categorical structure of NDs.

Moreover, since Cluster 2 displayed the most cognitive difficulties and included the highest number of individuals with formal NDs, it is interesting that this group was not also characterised by the weakest mental health profile. Research has shown that difficulties associated with neurodiverse conditions increase the risk for mental health problems, such as internalising (Baraskewich & McMorris, 2019). Thus, it would be expected that more severe cognitive profiles would co-occur with increased mental health difficulties. Whilst Cluster 1 was characterised by high emotional difficulty and moderate behavioural regulation, these dimensions did not effectively differentiate Cluster 1 from the other data-driven groups. Such results could reflect variability in mental health functioning, with some members of Cluster 1 displaying intact mental health (thus closer to Cluster 1), whilst others showed more severe difficulties (thus closer to Cluster 3), placing this cluster in the ‘middle ground’ relative to the other two groups. This heterogeneous mental health profile of Cluster 1 is aligned with the broad range of diagnostic labels captured by this group, as well as with literature suggesting that the mental health functioning of neurodivergent individuals varies widely (e.g., Obsuth et al., 2020; Shapiro et al., 2023). Such variability in the mental health profiles of individuals captured by this cluster may be related to the cognitive flexibility and processing speed differences that distinguish this group. Research suggests that cognitive flexibility may serve as a management tool for self-regulation and mental wellbeing, whilst processing speed has been suggested to play a ‘bridging’ role between cognition and emotion regulation strategies in adults (Goldin et al., 2025; Castro et al., 2025). Therefore, it is possible that individual differences in these cognitive domains may partly explain the inconsistent mental health presentations observed within this cluster. This aligns with the notion that neurodivergent profiles are not uniform, but shaped by the complex interplay between cognitive and mental health processes (e.g., D’Agati et al., 2019; Tajik-Parvinchi et al., 2021). Taken together, these results demonstrate how transdiagnostic approaches may provide a more meaningful framework for understanding the diversity of neurodivergent experiences, offering valuable insights into how cognitive and mental health processes may intersect in adulthood.

Cluster 3 could be interpreted as the ‘high functioning’ group despite significantly high hyperactivity and emotional difficulty, displaying the strongest cognitive profile amongst the data-driven groups. This profile was best defined by a combination of both dimensions, and characterised by strong cognitive flexibility, processing speed, language and working memory abilities. Cluster 3 also captured the highest number of individuals with ADHD (making up 25% of the cluster). Interestingly, the mental health and behavioural profile of this group aligns with the emotional and behavioural regulation difficulties characteristic of ADHD (e.g., Retz et al., 2012; APA, 2013), yet the cognitive profile diverges considerably from the well-established cognitive weaknesses associated with this condition (e.g., Onandia-Hinchado et al., 2021; Kofler et al., 2010; Holmes et al., 2010; Leib et al., 2021). This cluster may therefore represent a ‘resilient’ subgroup of individuals with ADHD characterised by emotional and behavioural difficulties, with strong cognitive functioning. These results suggest that despite real-world challenges, some individuals with ADHD may demonstrate strong task-based performance in cognition, providing further evidence for the heterogeneous nature of NDs.

The profile of Cluster 3 is important to consider in the context of future support strategies. For instance, cognitive control training has been shown to improve mental health outcomes (Koster et al., 2017), and many interventions, such as cognitive behavioural therapy, harness cognitive strategies to effectively target a range of mental health problems (e.g., Tolin, 2010). The present findings have therefore highlighted several cognitive areas which may serve as potential targets for such individualised support. Since the profile of Cluster 3 comprised individuals with various diagnostic labels (including autism and dyslexia), as well as those with suspected yet undiagnosed NDs, such support strategies may have transdiagnostic relevance and potentially benefit individuals based on cognitive/behavioural presentations, as opposed to being accessible solely on the basis of a diagnostic label. These findings illustrate the value of transdiagnostic, dimensional approaches for identifying meaningful targets for support that may be relevant across a wide range of profiles, and provide a foundation for developing support strategies that are both evidence-based and more responsive to individual needs.

The transdiagnostic dimensional design of the study directly addressed several shortcomings of categorical research. First, through the employment of a more relaxed approach to recruitment, the present study addressed the limitations of ‘case-control’ study designs, which have been shown to lead to a loss of important information regarding the complexity associated with NDs (Astle et al., 2022). To this end, the study’s inclusive sample captured a broad spectrum of abilities, from typical to atypical functioning, making it highly representative of the heterogeneous neurodiverse population, and allowing for the exploration of mechanisms that may contribute to such heterogeneity. Additionally, through focusing on neurodivergent adults specifically, the study addressed the current lack of research into the complexity of NDs in adulthood.

Second, the combination of dimensional cognitive and mental health assessments complemented the transdiagnostic nature of the study, and addressed some of the limitations associated with ‘core deficit’ accounts of diagnostic categories (Astle & Fletcher-Watson, 2020). This approach allowed for the exploration of strengths and weaknesses across several domains of functioning, therefore providing more valuable information than that offered by ‘gold standard’ assessments, which typically focus on the atypical end of the spectrum and may not be suitable for differentiating between individuals with milder difficulties (Alexander et al., 2020). Another strength of this broader approach to assessment is that it highlighted interactions between both cognitive and mental health processes that may reflect areas of need relevant across a wide range of NDs, providing an even more comprehensive understanding than cognition or mental health alone.

Third, the data-driven analytical approach employed was well-suited for this study’s transdiagnostic sample and the dimensional assessments administered, and enabled for the uncovering of valuable insights from such complex, multivariate data. The combination of data reduction methods allowed for a more comprehensive understanding of the underlying structure of the dataset and the nuanced between-variable interactions contributing to this structure. This multi-method approach enhanced the interpretability of the findings, through enabling the identification of meaningful patterns, which may be overlooked when adopting more traditional univariate or bivariate approaches that often accompany categorical or ‘case-control’ study designs (Astle et al., 2022). Importantly, the employment of data-driven methods offered the opportunity to explore the present dataset in a hypothesis-generating rather than hypothesis-confirming manner, which is particularly valuable in the context of neurodiversity research, where novel data-driven insights are crucial for advancing theoretical models. This allowed for the reconceptualisation of neurodiversity as a multidimensional space, in which naturally occurring transdiagnostic groups were identified, without confinement to predefined diagnostic groups, allowing for a shift away from reinforcing the categorical structure of NDs.

## Conclusion

The findings of this study suggest that neurodivergent cognitive and mental health profiles exist independently of traditional diagnostic categories, and that diagnosis alone may not comprehensively capture the complexity of individual functioning. This study therefore adds to the growing body of empirical evidence advocating for data-driven approaches that better capture real-world diversity (e.g., Astle et al., 2022).

The present results underscore the importance of assessment and support practices that extend beyond diagnostic labels to include more comprehensive profiling of cognitive and emotional/behavioural strengths and weaknesses. For instance, some neurodivergent adults may benefit from targeted support in language and working memory, even in the absence of a formal dyslexia (or other SLD) diagnosis. Other individuals with more complex profiles (e.g., co-occurring conditions) may benefit from support targeting cognitive flexibility and processing speed as these domains may underpin both cognitive and mental health difficulties, and/or be relevant to a broad range of NDs. In contrast, individuals with strong cognitive profiles but marked mental health/behavioural difficulties may benefit from strengths-based approaches that harness preserved abilities to improve everyday functioning.

However, further research is needed to build on and enhance the translational impact of the present findings, and to extend current transdiagnostic dimensional frameworks. Future transdiagnostic research should continue to yield data-driven accounts of NDs and employ broader, dimensional assessments in order to establish the cognitive and mental health processes that contribute to individual variation and overlaps across NDs. Future studies should ensure sufficient sample sizes for detecting meaningful data-driven profiles, and efforts should be made to recruit participants from a variety of sources in order to increase the representativeness of the sample beyond convenience-based populations. This will be important for establishing the generalisability of data-driven profiles across the broader neurodiverse community. Additionally, future research should seek to explore alternative dimensionality reduction approaches. For instance, hierarchical clustering may provide the opportunity to give more weight to theoretically meaningful constructs that contribute less to overall variance, allowing for the formation of more balanced, data-driven groupings (e.g., Everitt et al., 2011); while network analysis could help to uncover patterns of between-variable interactions that may be overlooked in the process of clustering, allowing for the identification of processes that may act as key mediators or ‘bridging’ mechanisms between different parts of the network (Borsboom et al., 2021; Mareva et al., 2019). Incorporating these techniques in well-powered studies could provide further insights into the complex interplay between cognition and mental health in NDs, and validate the present findings.

## Data Availability

All data produced in the present study are available upon reasonable request to the authors.

## Acknowledgements

This work was supported by a PhD studentship (Project Reference: 2711931) awarded to KM with partial contribution from the UK Research and Innovation (UKRI) Engineering and Physical Sciences Research Council

The authors would like to thank Katharine Chisholm, Sarah Carrington and Jane Waite for their feedback on drafts of this manuscript. We also wish to thank all participants who have taken part in this study, and all organisations who have supported us with the recruitment of participants, including Aston University, Dyspraxia Foundation, UK Adult ADHD Network, Embracing Complexity, All Age Autism, The Recovery College (Black Country Healthcare NHS Foundation Trust), ADHD Adult UK, and the Cambridge Autism Research Database.

